# Evaluation of operational characteristics and performance of HIV rapid diagnostic tests (RDTs): Systematic review and meta-analysis of the literature from 2012 to 2020

**DOI:** 10.1101/2022.05.11.22274936

**Authors:** Leonard Kingwara, Nancy Bowen, Nuttada Panpradist, Peter Lokamar, Vera Morangi, Rukia Sarah Madada, Emmanuel Nyakeriga, Christabel Awuor, Jonah Onentia, Sarah Masyuko, Rose Wafula, John Ndemi Kiiru

## Abstract

**Background:** Most countries have rolled out HIV Rapid diagnostic tests (RDTs) due to their significant advantages over laboratory-based serological testing. These advantages are lower cost, ease of use, interpretation speed, and relatively high acceptability; the HIV diagnostic landscape has evolved fast, and newer technologies have been developed and deployed. Given the many options available, selecting an HIV rapid diagnostic test for a particular clinical program, self-test, or research setting can be daunting without the precise knowledge of their performance characteristics. This systematic review and meta-analysis aimed to compare the field diagnostic performance of available HIV rapid test kits, cost-effectiveness, ease of use, and acceptability.

**Methods:** PubMed and Web of Science were searched for publications on rapid HIV tests using blood specimens. We then performed a meta-analysis and systematic analysis to quantitatively and qualitatively evaluate the diagnostic performance of rapid HIV tests compared with the western blot (WB), enzyme-linked immunosorbent assay (ELISA), or an HIV diagnostic algorithm in terms of pooled sensitivity, specificity, area under the summary receiver operating characteristic (SROC) curve, and diagnostic odds ratio (DOR).

**Results:** The meta-analysis for the diagnostic test included 26 studies for diagnostic accuracy, while the qualitative analysis included 15 studies. On average, the RDT sensitivities were 99%; [95% CI=0.99-100%], while specificity was optimal at 100%; [95% CI=99%-100%]. The diagnostic odds ratio estimates that a single test performed better than a dual test: dual test DOR=44612.33 and single test DOR=14323.1. The impact of unobserved heterogeneity using the quantity I^2^ for sensitivity was 99.47%, while that for specificity was 99.96, indicating significant heterogeneity and justifying stratified analysis of the selected studies. The diagnostic test from Unigold had the best-pooled sensitivity, specificity, and diagnostic odds ratio at 99%, 99.35%, and 2896.667, respectively. Qualitative data indicate shorter time to results is preferred by both the clients and health care providers.

**Conclusion:** The average of the RDT sensitivities for diagnostic accuracy were 99% (95% CI=0.99-100%), while specificity was optimal at 100%; 95% CI=99-100. The diagnostic odds ratio was DOR=44612 (95% CI=14323-138954), thus indicating better RDT test performance. The performance of single test kits in HIV diagnosis was better than those for dual tests.

## Background and introduction

Rapid diagnostic tests (RDTs) have significant advantages over laboratory-based serological testing[1], including lower cost, ease of use, interpretation speed, and relatively high acceptability[2]. A systematic review by Olugbenga *et al*. pointed to a quick turn-around time of 20-60 minutes, eliminating the need for a second visit to the medical facilities to get test results[3].

Particularly, RDTs for HIV serology assays that detect the presence of HIV-1/2 antibodies had higher sensitivity and specificity than those of other infectious diseases[3]. RDTs have proven instrumental in increased HIV testing, allowing testing to be performed even by non-laboratory professionals in both communities and facilities (including sites with limited infrastructure, e.g., electricity)[4], [5]. Such facilities register and process low numbers of specimens daily[5]. Thus, the ease of use, cost-effectiveness, acceptability, and transportability of RDTs have resulted in higher rates of HIV diagnoses globally[4].

Additionally, the availability of rapid diagnostic kits (RDKs) has assisted in moving the proportion of those aware of their HIV positivity status from an estimated 10% in sub-Saharan Africa to 76% as of 2016 in East and Southern Africa [4]. Most countries are approaching the 90% mark of people living with HIV (PLHIV) who know their status as documented by a joint United Nations Programme (UNP) on HIV/AIDS (UNAIDS) benchmark[5].

Most of the systematic reviews have limited their scope to the performance characteristics of the test Kits. We have assessed the expanding success of the RDT program using existing data to assess three major outcomes of HIV self-testing (HIVST); cost-effectiveness, ease of use, and acceptability to drive accountability. Therefore, the objective of this systematic review was to evaluate the performance and operation of RDTs to inform on the global HIV response.

## Materials and methods

### Eligibility criteria

We followed the preferred reporting items for systematic reviews and meta-analysis guidelines (PRISMA) for this systematic review[6]. We have included **s**tudies evaluating commercially available rapid diagnostic test kits for HIV in laboratory or field settings. The studies includedwere for all populations in any geographic location. The primary outcome was diagnostic test accuracy (i.e., sensitivity, specificity, positive predictive value, negative predictive values). Secondary outcomes were cost-effectiveness, usability, ease of use, and acceptability. The eligible studies for inclusion were evaluation studies, cost-effectiveness studies, and usability and acceptability studies. Studies were included for the meta-analysis of diagnostic accuracy if an acceptable reference standard for HIV test kit evaluation was used. This included either enzyme immunoassay (EIA), Western blot (WB), or an HIV diagnostic algorithm. Studies were excluded if they did not provide any information regarding RTDs in the context of accuracy or cost-effectiveness, usability, ease of use, and acceptability. We included studies regardless of sample size and regional location.

### Search terms and strategy

Medline, Embase, ScienceDirect, and Google Scholar were searched using Boolean operators to explore terms related to the following three concepts: HIV, rapid diagnostic test kits, and diagnosis. Only studies published in English were included. Studies published between January 2012 and August 2020 were included. The searches were rerun immediately before the final analyses to check for recent relevant literature. Additional records were identified by searching bibliographies of relevant publications.

### Data extraction

Titles and abstracts were scrutinized to assess relevance. For the meta-analysis of diagnostic accuracy, the data extracted included study title, country, test(s) evaluated, laboratory or field evaluation (and if so, sample type used), the population studied, and laboratory evaluations, whether fresh blood, saliva, or archived specimens were used. The total number of participants/samples used, prevalence (%), reference standard test, and the number of true positives, false positives, false negatives, and true negatives were all determined if they were not provided in the review publications. Seven reviewers independently extracted data from the included studies, and consensus resolved disagreements. To evaluate the methodology of the included studies, updated standards for reporting diagnostic accuracy studies (STARD) checklist was adopted [7]. Additionally, the quality assessment of diagnostic accuracy studies (QUADAS-2) checklist was adopted to appraise the included evaluation studies [8] critically.

### Data analysis plan

Test indicators outlined in **Table 1** below were analyzed. Forest plots (*Midas* package) and summary receiver operating characteristic (SROC) curves were constructed using Stata software v16.1(Stata Corp LP, College Station, Texas, USA). Additionally, the impact of unobserved heterogeneity using the quantity I^2^ was assessed. Considering that rigorous statistical analysis requires hierarchical (multilevel) models that respect the binomial data structure (hierarchical logistic regression), *Metandi* Stata package was used to facilitate the fitting of such models. The commands display the results in two alternative parameterizations and produce a customizable plot[9]. The diagnostic odds ratio (DOR) as a measure of test performance combines the strengths of sensitivity and specificity, as independent prevalence indicators, with the advantage of accuracy as a single indicator[10]. A DOR >1 indicates better test performance for the likelihood of deducting disease amongst a healthy population.

**Table 1.**
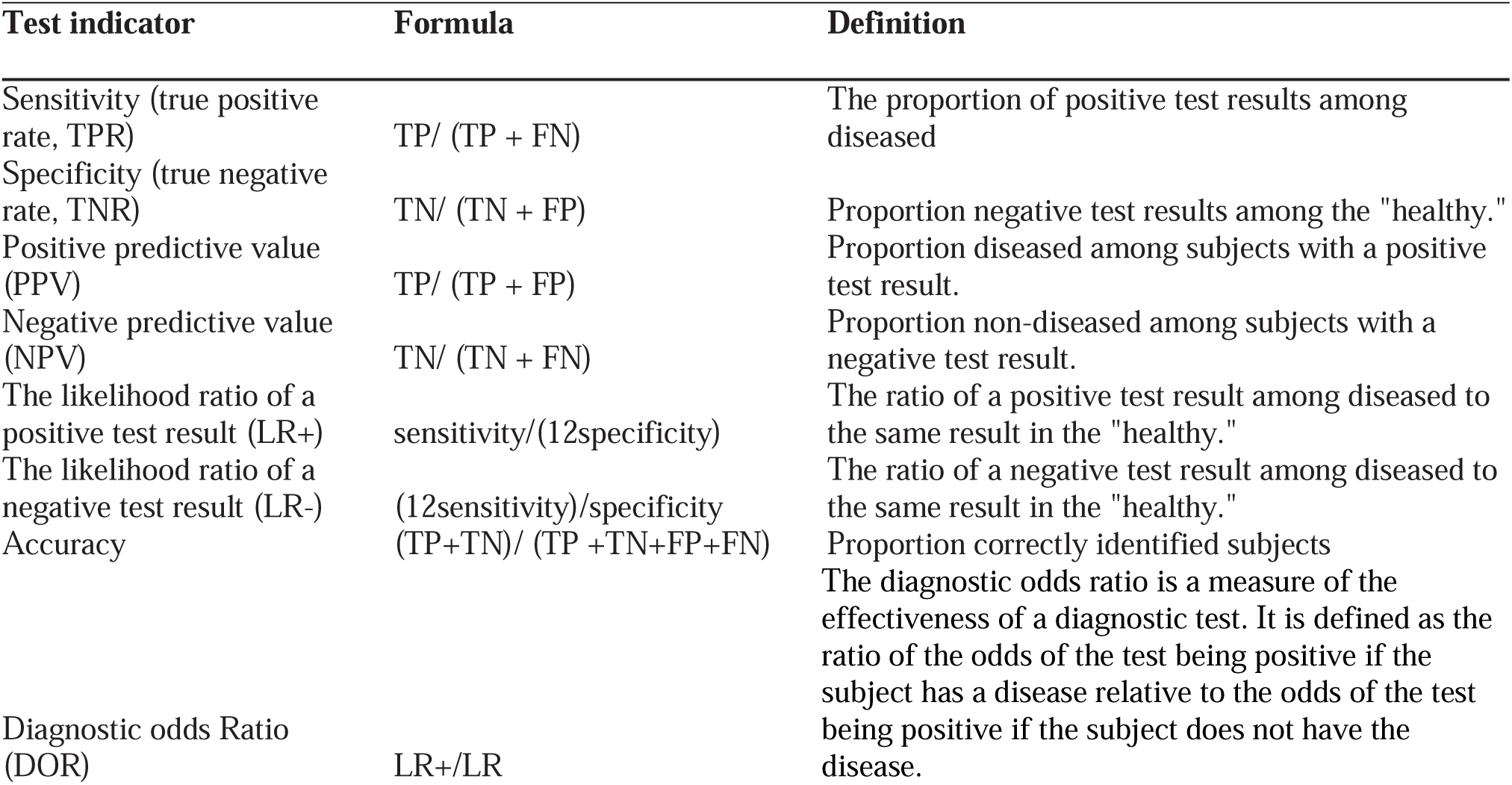
Definition of indicators used in meta-analysis of test performance.

| Test indicator | Formula | Definition |
| --- | --- | --- |
| Sensitivity (true positive rate, TPR) | $TP / (TP + FN)$ | The proportion of positive test results among diseased |
| Specificity (true negative rate, TNR) | $TN / (TN + FP)$ | Proportion negative test results among the "healthy." |
| Positive predictive value (PPV) | $TP / (TP + FP)$ | Proportion diseased among subjects with a positive test result. |
| Negative predictive value (NPV) | $TN / (TN + FN)$ | Proportion non-diseased among subjects with a negative test result. |
| The likelihood ratio of a positive test result (LR+) | $sensitivity / (1 - specificity)$ | The ratio of a positive test result among diseased to the same result in the "healthy." |
| The likelihood ratio of a negative test result (LR-) | $(1 - sensitivity) / specificity$ | The ratio of a negative test result among diseased to the same result in the "healthy." |
| Accuracy | $(TP + TN) / (TP + TN + FP + FN)$ | Proportion correctly identified subjects |
| Diagnostic odds Ratio (DOR) | LR+ / LR- | The diagnostic odds ratio is a measure of the effectiveness of a diagnostic test. It is defined as the ratio of the odds of the test being positive if the subject has a disease relative to the odds of the test being positive if the subject does not have the disease. |

### Study characteristics

The review included the analysis of the diagnostic tests for a single rapid HIV test, a dual HIV/syphilis test, and a triple HIV/syphilis/HCV and evaluated the HIV diagnostic component of the rapid HIV test device. However, there were limited data to evaluate dual or triple HIV rapid test kits.

Diagnostic accuracy publications evaluated the performance of INSTI™ HIV-1 Antibody Test (HIV-1), DPP^®^ HIV-HCV Screen Assay Oral Test (HIV-HCV Screen Oral), DPP^®^ HIV-HCV-Syphilis Screen Assay Blood Test (HIV-HCV-Syphilis Blood), Multiplo Rapid HIV/HCV/HBV Antibody Test (HIV/HCV/HBV), DPP^®^ HIV 1/2 Screen Assay Blood Test (HIV 1/2 Screen Blood), Alere Determine HIV 1&2, DPP Rapid Test HIV 1/2, DS Rapid Test HIV, Interknit HIV 1&2, HIV 1/2/O Tri-Line, Immuno-Rapido HIV 1&2, Imunocrom HIV 1/2, StatPak, Capillus, Hexagon HIV, First Response HIV1-2-O, SD Bioline HIV-1/2 3.0, HIV Tri-Dot + AG, Genie™ III HIV1/2, INSTI VIH1/2, Genie Fast™ HIV 1-2, SD Bioline HIV/SYPHILIS Duo, Vikia HIV1/2, OraQuick Advance Rapid HIV-1/2, Chembio DPP HIV 1/2 Assay, Chembio Sure Check HIV1/2 Assay, Precise HIV, and BCP HIV-1/-2. The qualitative studies evaluated the acceptability, ease of use, and cost-effectiveness of rapid diagnostic HIV test kits.

These were global studies ranging from countries in Africa, South America, Asia, and the USA to Europe. The populations studied included key populations (men who have sex with men, female sex workers (FSWs), sexual health clinic attendees), adults from the general population, children, archived samples, nursing and pharmacy students, and women in antenatal and post-partum clinics. One study was done among truck drivers in the field.

## Results

### Included and excluded studies

The meta-analysis for the diagnostic test included 26 studies for diagnostic accuracy, while the qualitative analysis included 15 studies, as shown in **Figure 1**. The sample sizes for each study ranged from 166 to 4,458. Apart from the rapid diagnostic tests, one self-test Kit (INSTI-test) that assessed diagnostic accuracy using blood samples of adults visiting a clinic in Kenya was included. The findings of this study indicated an accuracy of 97.9%, with a sensitivity of 98%, a specificity of 97.8%, a negative predictive value of 98.5%, and a positive predictive value of 97.9%. This study was included in the meta-analysis because it tested accuracy of the INSTI-test against enzyme-linked immunosorbent assay (ELISA) as the gold standard. The systematic review included a publication from Mugwanya and Pintye et al. on the feasibility and programmatic evaluation of integrating pre-exposure prophylaxis delivery in routine family planning Kenyan clinics (ref). While the results reported the accuracy of HIV testing, this study was excluded in the meta-analysis since it did not evaluate any RDT. Characteristics and results of studies evaluating the diagnostic test accuracy of dual HIV RDTs are in **Table 2a-2d**.

**Figure 1.**
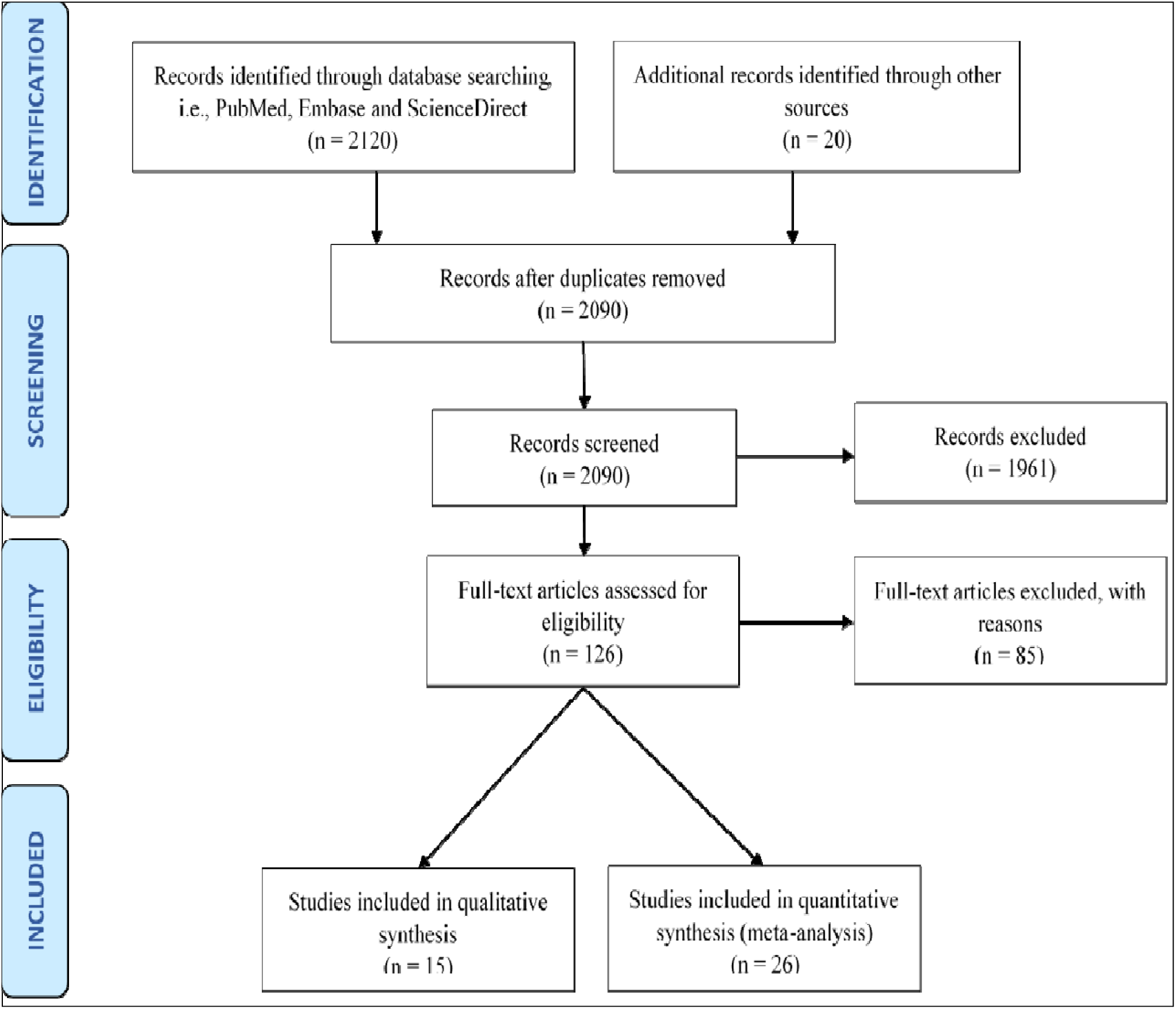
PRISMA diagram for the present systematic review.

**Figure 2.**
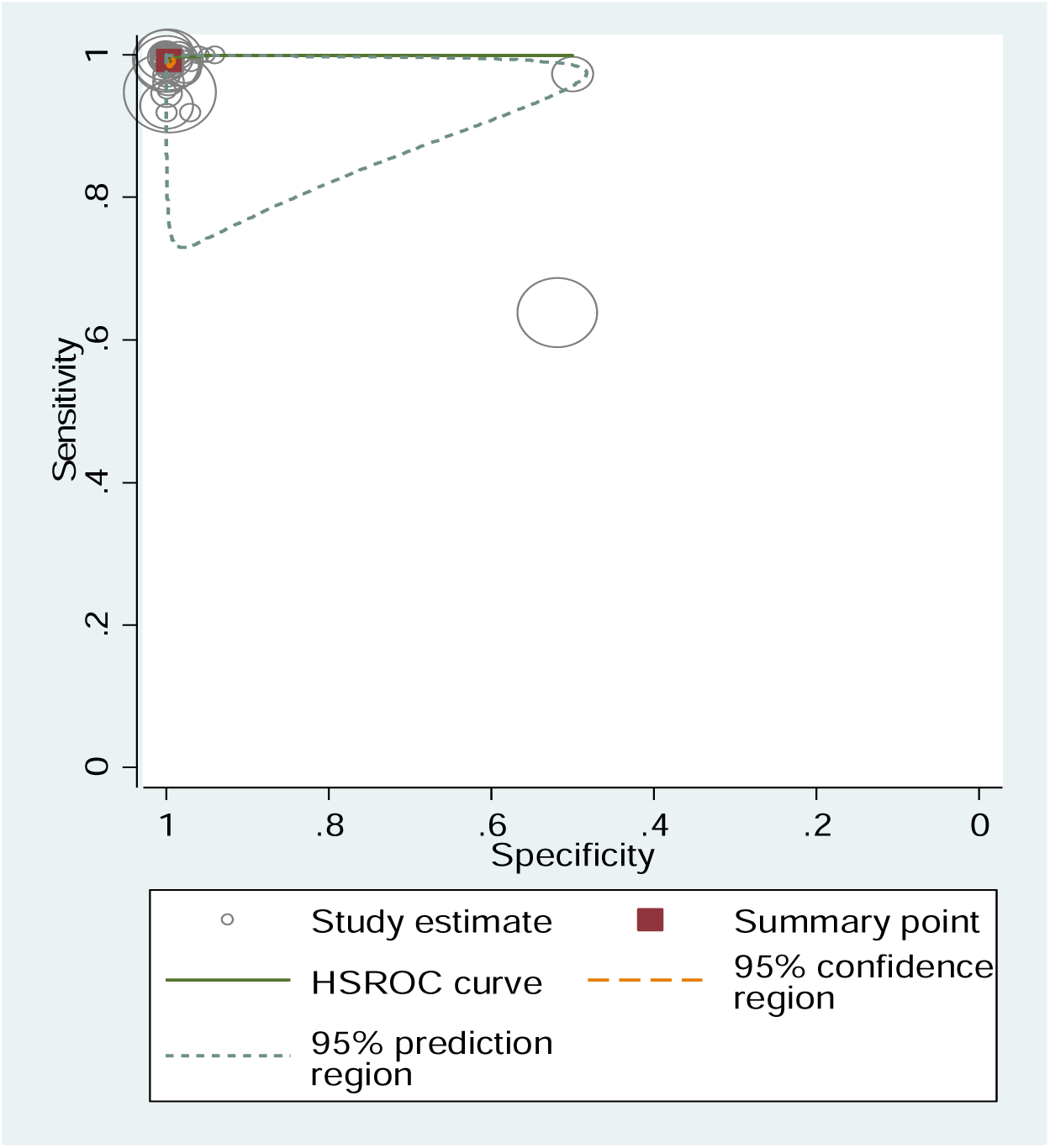
HSROC. Hierarchical summary ROC: ROC = receiver operating characteristic of the test Kits evaluated for quantitative analysis. The pooled sensitivity, specificity, and area under the SROC curve of these assays were 0.999, 0.991, and 1.00, respectively, which demonstrate that blood-based rapid HIV test has comparable accuracy to WB, RDT testing algorithms, or ELISA for HIV early therapy.

**Table 2a.** Characteristics and results of studies evaluating the diagnostic test accuracy of dual HIV RDTs included in the meta-analysis; Test site; Clinic.

| Author | Location | Population | Index test | Sample | Sample size | Reference test | TP | FN | FP | TN | Accuracy | Sens | Spec | NPV | PPV | Prev |
| --- | --- | --- | --- | --- | --- | --- | --- | --- | --- | --- | --- | --- | --- | --- | --- | --- |
| Choko [12] 2016 | Malawi | Adults | OraQuick | Saliva | 378 | Determine or Unigold | 175 | 5 | 0 | 190 | 365 | 97.2 | 100 | 97 | 100 | 47.6 |
| Choko [12] 2016 | Malawi | Adults | OraQuick | Blood | 378 | Determine or Unigold | 174 | 3 | 1 | 186 | 360 | 99.5 | 99.5 | 98 | 99.4 | 46.8 |
| Maheswaran [13] 2016 | South Africa | Adults | OraQuick | Saliva | 2198 | Determine or Unigold | 323 | 4 | 0 | 1860 | 327 | 98.7 | 100 | 100 | 100 | 14.8 |
| Nangendo [14] 2017 | Uganda | Adults | OraQuick | Saliva | 440 | N/A | 65 | 0 | 0 | 375 | 440 | 100 | 100 | 100 | 100 | 14.8 |
| Nkenfou [15] 2013 | Cameroon | Key populations | OraQuick | Blood | 1520 | Determine or Unigold | 13 | 1 | 1 | 1505 | 99.9 | 93 | 99.9 | 100 | 92.9 | 1 |
| Aspard [16] 2014 | France | General populations | OraQuick | Saliva/Whole blood | 179 | ChemBio Sure Check HIV | n/a | n/a | n/a | n/a | n/a | 87.2 | 98 | n/a | n/a | n/a |
| Aspard [16] 2014 | France | General populations | DPP-HIV | Saliva/Whole blood | 179 | ChemBio Sure Check HIV1/2 Assay | n/a | n/a | n/a | n/a | n/a | 88.3 | 100 | n/a | n/a | n/a |
| Aspard [16] 2014 | France | General populations | DPP-HIV | Saliva/Whole blood | 179 | ChemBio Sure Check HIV1/2 Assay | n/a | n/a | n/a | n/a | n/a | 100 | 100 | n/a | n/a | n/a |
| Obachukwu [17] 2015 | Nigeria | Women | Determine | Serum | 166 | ELISA | 19 | 1 | 0 | 146 | 99.4 | 95 | 100 | 99 | 100 | 12.05 |
| Juntharasmalai [18] 2015 | Thailand | Key populations | OraQuick | Saliva/Whole blood | 3678 | ELISA | 436 | 2 | 10 | n/a | n/a | 99.5 | 99.7 | 100 | 97.8 | n/a |
| Olugbenga [17] 2018 | Nigeria | Women | SD Bioline/Syphilis | Serum | 4551 | ELISA |  | n/a | n/a | n/a | n/a | 85.8 | 99.5 | 0.9 | 0.15 | n/a |
| Rapeeporn [19] 2018 | Thailand | Adults | SD Bioline/Syphilis | Blood | 1680 | MTs using Serodia HIV 1/2 | 252 | 143 | 1428 | 1537 | n/a | 0.638 | 0.52 | 0.9 | 0.15 | 23.51 |
| Wawira [11] 2018 | Kenya | Adults | INSTI-test | Blood | 470 | ELISA | 196 | 4 | 6 | 264 | 97.9 | 98 | 97.8 | 99 | 97 | 42.55 |

Total Positives (TP), Total Negatives (TN), False Positives (FP), and False Negatives (FN)
Key populations: men who have sex with men, female sex workers (FSWs), and sexual health clinic attendees

**Table 2b.** Characteristics and results of studies evaluating the diagnostic test accuracy of dual HIV RDTs included in the meta-analysis; Test site; Field.

| Author | Location | Population | Index test | Sample | Sample size | Reference test | TP | FN | FP | TN | Accuracy | Sens | Spec | NPV | PPV | Prev |
| --- | --- | --- | --- | --- | --- | --- | --- | --- | --- | --- | --- | --- | --- | --- | --- | --- |
| Belete [20] 2019 | Ethiopia | Adults | OraQuick | Saliva | 400 | Determine or Unigold | 200 | 1 | 0 | 200 | 399 | 99.5 | 100 | 99.5 | 100 | 50 |
| Dziva 2019[21] | Kenya/<br>Zimbabwe | Children | OraQuick | Saliva | 1776 | Determine or Unigold | 71 | 0 | 0 | 1705 | 1776 | 100 | 100 | 100 | 100 | 4 |
| Fishers [22]2019 | USA | Adults | DPP-HIV | Saliva | 1029 | ELISA | n/a | n/a | n/a | n/a | n/a | 97.2 | 99.5 | n/a | n/a | n/a |
| Fishers [22]2019 | USA | Adults | DPP-HIV/HCV | Saliva | 1029 | ELISA | n/a | n/a | n/a | n/a | n/a | 97.9 | 99.5 | n/a | n/a | n/a |
| Fishers 2019[22] | USA | Adults | Determine | Blood | 1029 | ELISA | n/a | n/a | n/a | n/a | n/a | 94.6 | 99.9 | n/a | n/a | n/a |
| Fishers [22]2019 | USA | Adults | DPP-HIV/HCV/Syphilis | Blood | 1029 | ELISA | n/a | n/a | n/a | n/a | n/a | 100 | 99.9 | n/a | n/a | n/a |
| Fishers [22]2019 | USA | Adults | INSTI-Test | Blood | 1029 | ELISA | n/a | n/a | n/a | n/a | n/a | 95.5 | 100 | n/a | n/a | n/a |
| Fishers [22]2019 | USA | Adults | HIV/HCV/HBV | Blood | 1029 | ELISA | n/a | n/a | n/a | n/a | n/a | 94 | 100 | n/a | n/a | n/a |
| Fishers [22]2019 | USA | Adults | DPP-HIV/HCV | Blood | 1029 | ELISA | n/a | n/a | n/a | n/a | n/a | 98 | 99.4 | n/a | n/a | n/a |
| Fishers [22]2019 | USA | Adults | DPP-HIV/HCV | Blood | 1029 | ELISA | n/a | n/a | n/a | n/a | n/a | 100 | 99.7 | n/a | n/a | n/a |
| Fishers [22]2019 | USA | Adults | HIV/Syphilis test | Blood | 1029 | ELISA | n/a | n/a | n/a | n/a | n/a | 97 | 99.8 | n/a | n/a | n/a |
| Fishers [22]2019 | USA | Adults | DPP-HIV/HCV | Blood | 1029 | ELISA | n/a | n/a | n/a | n/a | n/a | 91.1 | 100 | n/a | n/a | n/a |
| Zachary [23]2012 | Zambia | Key populations | OraQuick | Saliva/<br>Whole blood | 4458 | Determine or Unigold | 721 | 40 | 15 | 3682 | 98.8 | 94.7 | 99.6 | 98.9 | 98 | 17.1 |
| Galiwango [24]2013 | Uganda | Key populations | Determine | Blood | 859 | EIA-WB | 832 | 23 | 2 | 2 | 97.1 | 97.3 | 99.9 | 98.6 | 99.8 | 99.5 |
| Galiwango [24]2013 | Uganda | Key populations | StatPak | Blood | 2540 | EIA-WB | 869 | 5 | 4 | 1662 | 99.6 | 99.6 | 99.7 | 99.8 | 99.4 | 34.2 |
| Asimwe [25]2014 | Uganda | Key populations | OraQuick | Saliva/<br>Whole blood | 246 | Determine<br>or Unigold | n/a | n/a | n/a | n/a | n/a | n/a | n/a | n/a | n/a | n/a |
| Madisa [26]2015 | Botswana | Key populations | KHB/Unigold | Blood | 875 | ELISA | 474 | 9 | 7 | 385 | 98.2 | 98.1 | 98.2 | 97.7 | 98.5 | 55.2 |
| Madisa [26]2015 | Botswana | Key populations | KHB/Unigold | Blood | 297 | ELISA | 31 | 0 | 4 | 262 | 88.2 | 100 | 98.5 | 100 | 88.6 | 10.4 |

Total Positives (TP), Total Negatives (TN), False Positives (FP), and False, Negatives (FN)
Key populations: men who have sex with men, female sex workers (FSWs), and sexual health clinic attendees

**Table 2c.**
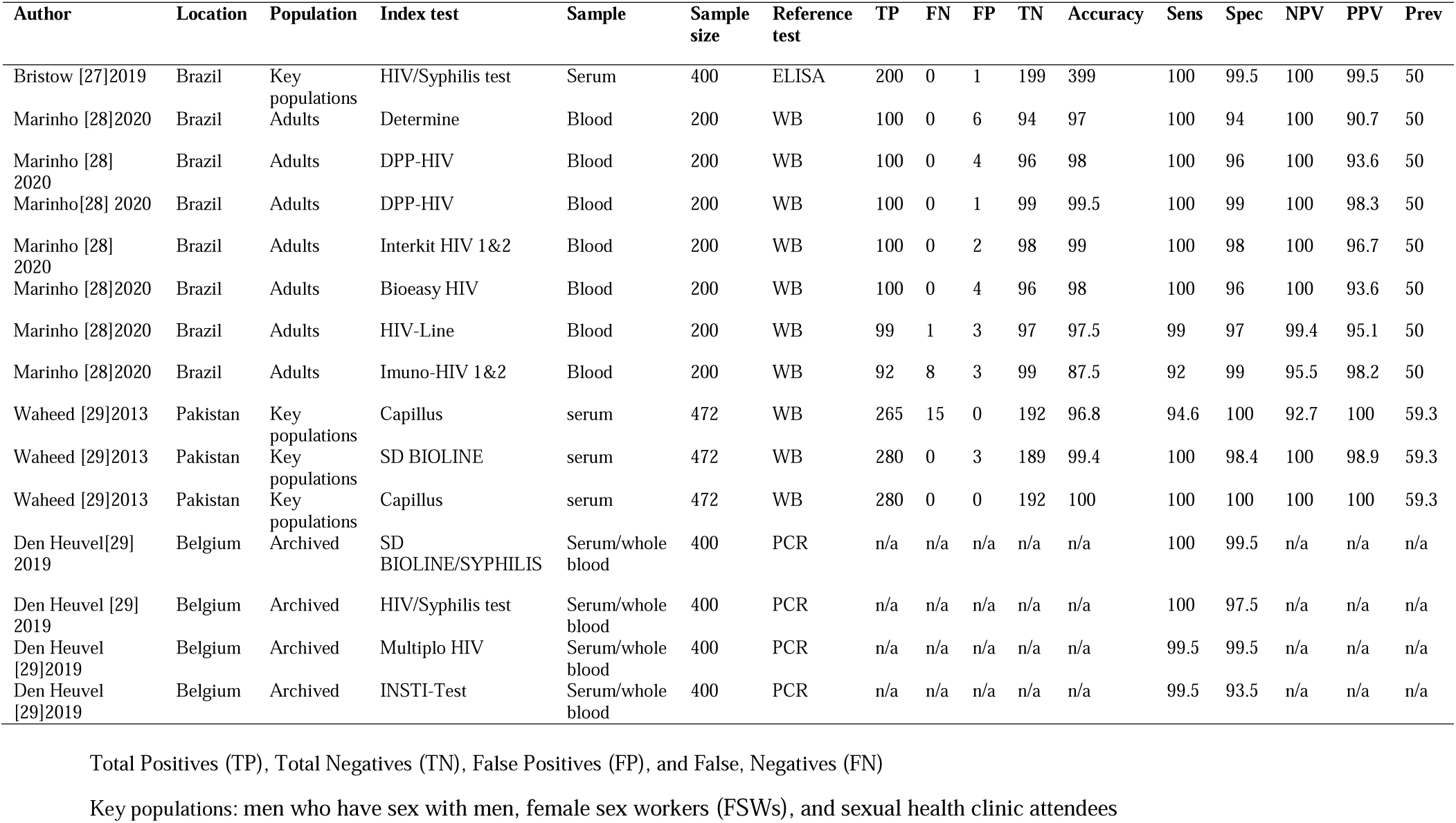
Characteristics and results of studies evaluating the diagnostic test accuracy of dual HIV RDTs that were in the meta-analysis; Test site; Laboratory.

| Author | Location | Population | Index test | Sample | Sample size | Reference test | TP | FN | FP | TN | Accuracy | Sens | Spec | NPV | PPV | Prev |
| --- | --- | --- | --- | --- | --- | --- | --- | --- | --- | --- | --- | --- | --- | --- | --- | --- |
| Bristow [27]2019 | Brazil | Key populations | HIV/Syphilis test | Serum | 400 | ELISA | 200 | 0 | 1 | 199 | 399 | 100 | 99.5 | 100 | 99.5 | 50 |
| Marinho [28]2020 | Brazil | Adults | Determine | Blood | 200 | WB | 100 | 0 | 6 | 94 | 97 | 100 | 94 | 100 | 90.7 | 50 |
| Marinho [28] 2020 | Brazil | Adults | DPP-HIV | Blood | 200 | WB | 100 | 0 | 4 | 96 | 98 | 100 | 96 | 100 | 93.6 | 50 |
| Marinho[28] 2020 | Brazil | Adults | DPP-HIV | Blood | 200 | WB | 100 | 0 | 1 | 99 | 99.5 | 100 | 99 | 100 | 98.3 | 50 |
| Marinho [28] 2020 | Brazil | Adults | Interkit HIV 1&2 | Blood | 200 | WB | 100 | 0 | 2 | 98 | 99 | 100 | 98 | 100 | 96.7 | 50 |
| Marinho [28]2020 | Brazil | Adults | Bioeasy HIV | Blood | 200 | WB | 100 | 0 | 4 | 96 | 98 | 100 | 96 | 100 | 93.6 | 50 |
| Marinho [28]2020 | Brazil | Adults | HIV-Line | Blood | 200 | WB | 99 | 1 | 3 | 97 | 97.5 | 99 | 97 | 99.4 | 95.1 | 50 |
| Marinho [28]2020 | Brazil | Adults | Imuno-HIV 1&2 | Blood | 200 | WB | 92 | 8 | 3 | 99 | 87.5 | 92 | 99 | 95.5 | 98.2 | 50 |
| Waheed [29]2013 | Pakistan | Key populations | Capillus | serum | 472 | WB | 265 | 15 | 0 | 192 | 96.8 | 94.6 | 100 | 92.7 | 100 | 59.3 |
| Waheed [29]2013 | Pakistan | Key populations | SD BIOLINE | serum | 472 | WB | 280 | 0 | 3 | 189 | 99.4 | 100 | 98.4 | 100 | 98.9 | 59.3 |
| Waheed [29]2013 | Pakistan | Key populations | Capillus | serum | 472 | WB | 280 | 0 | 0 | 192 | 100 | 100 | 100 | 100 | 100 | 59.3 |
| Den Heuvel[29] 2019 | Belgium | Archived | SD BIOLINE/SYPHILIS | Serum/whole blood | 400 | PCR | n/a | n/a | n/a | n/a | n/a | 100 | 99.5 | n/a | n/a | n/a |
| Den Heuvel [29] 2019 | Belgium | Archived | HIV/Syphilis test | Serum/whole blood | 400 | PCR | n/a | n/a | n/a | n/a | n/a | 100 | 97.5 | n/a | n/a | n/a |
| Den Heuvel [29]2019 | Belgium | Archived | Multiplo HIV | Serum/whole blood | 400 | PCR | n/a | n/a | n/a | n/a | n/a | 99.5 | 99.5 | n/a | n/a | n/a |
| Den Heuvel [29]2019 | Belgium | Archived | INSTI-Test | Serum/whole blood | 400 | PCR | n/a | n/a | n/a | n/a | n/a | 99.5 | 93.5 | n/a | n/a | n/a |

Total Positives (TP), Total Negatives (TN), False Positives (FP), and False, Negatives (FN)
Key populations: men who have sex with men, female sex workers (FSWs), and sexual health clinic attendees

**Table 2d:**
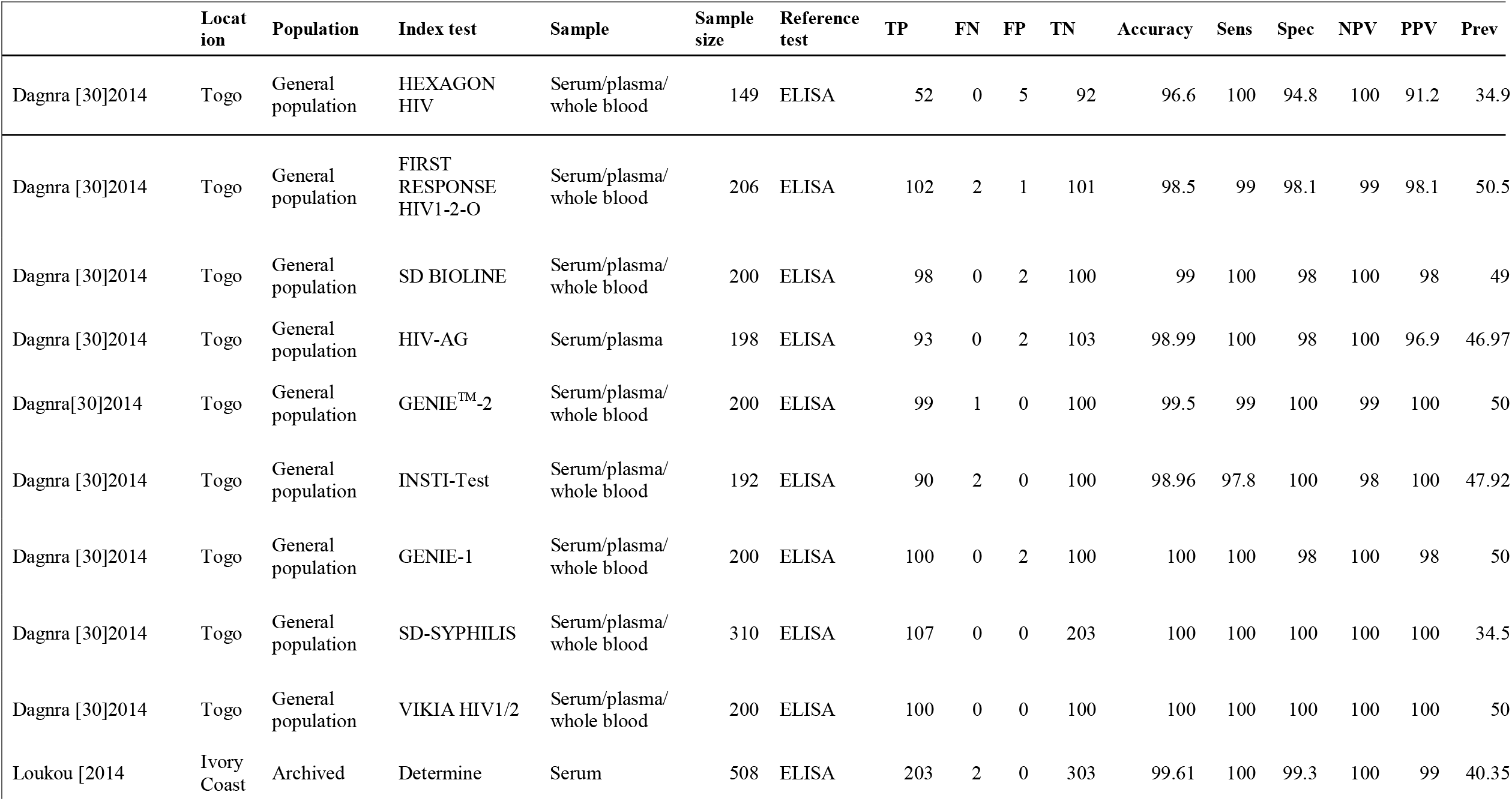
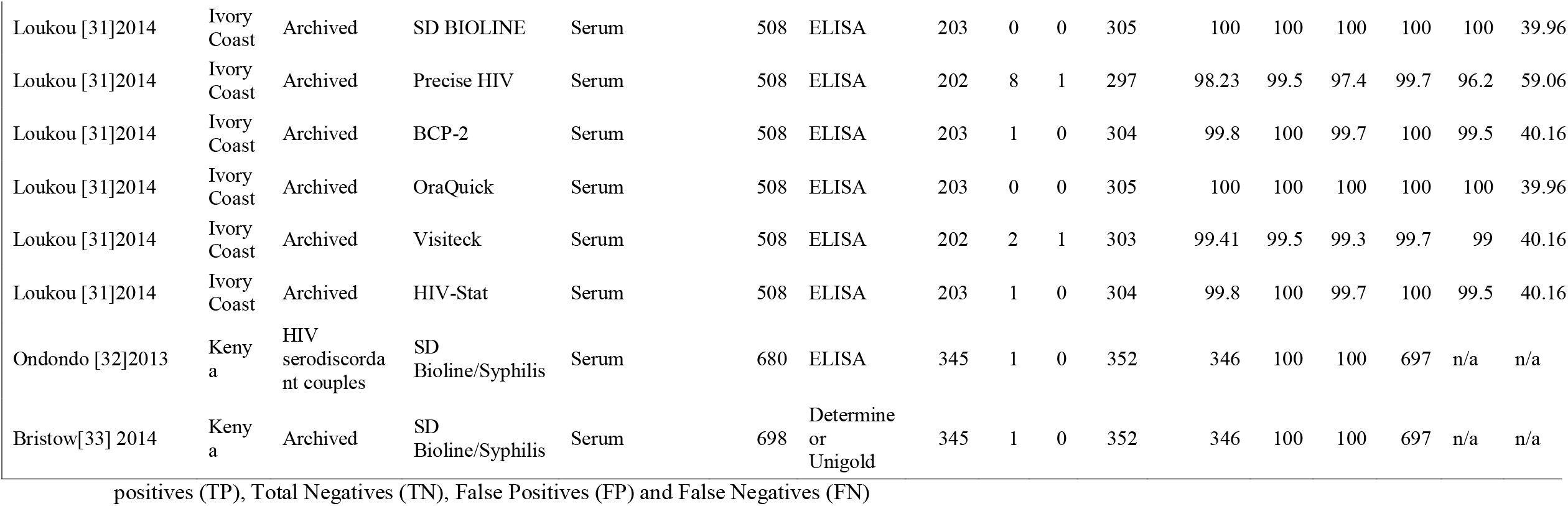
Characteristics and results of studies evaluating the diagnostic test accuracy of dual HIV RDTs that were included in the meta-analysis; Test site; Laboratory.

|  | Location | Population | Index test | Sample | Sample size | Reference test | TP | FN | FP | TN | Accuracy | Sens | Spec | NPV | PPV | Prev |
| --- | --- | --- | --- | --- | --- | --- | --- | --- | --- | --- | --- | --- | --- | --- | --- | --- |
| Dagnra [30]2014 | Togo | General population | HEXAGON HIV | Serum/plasma/whole blood | 149 | ELISA | 52 | 0 | 5 | 92 | 96.6 | 100 | 94.8 | 100 | 91.2 | 34.9 |
| Dagnra [30]2014 | Togo | General population | FIRST RESPONSE HIV1-2-O | Serum/plasma/whole blood | 206 | ELISA | 102 | 2 | 1 | 101 | 98.5 | 99 | 98.1 | 99 | 98.1 | 50.5 |
| Dagnra [30]2014 | Togo | General population | SD BIOLINE | Serum/plasma/whole blood | 200 | ELISA | 98 | 0 | 2 | 100 | 99 | 100 | 98 | 100 | 98 | 49.7 |
| Dagnra [30]2014 | Togo | General population | HIV-AG | Serum/plasma | 198 | ELISA | 93 | 0 | 2 | 103 | 98.99 | 100 | 98 | 100 | 96.9 | 46.97 |
| Dagnra[30]2014 | Togo | General population | GENIE™-2 | Serum/plasma/whole blood | 200 | ELISA | 99 | 1 | 0 | 100 | 99.5 | 99 | 100 | 99 | 100 | 50.0 |
| Dagnra [30]2014 | Togo | General population | INSTI-Test | Serum/plasma/whole blood | 192 | ELISA | 90 | 2 | 0 | 100 | 98.96 | 97.8 | 100 | 98 | 100 | 47.92 |
| Dagnra [30]2014 | Togo | General population | GENIE-1 | Serum/plasma/whole blood | 200 | ELISA | 100 | 0 | 2 | 100 | 100 | 100 | 98 | 100 | 98 | 50.0 |
| Dagnra [30]2014 | Togo | General population | SD-SYPHILIS | Serum/plasma/whole blood | 310 | ELISA | 107 | 0 | 0 | 203 | 100 | 100 | 100 | 100 | 100 | 34.5 |
| Dagnra [30]2014 | Togo | General population | VIKIA HIV1/2 | Serum/plasma/whole blood | 200 | ELISA | 100 | 0 | 0 | 100 | 100 | 100 | 100 | 100 | 100 | 50.0 |
| Loukou [2014 | Ivory Coast | Archived | Determine | Serum | 508 | ELISA | 203 | 2 | 0 | 303 | 99.61 | 100 | 99.3 | 100 | 99 | 40.35 |
| Loukou [31]2014 | Ivory Coast | Archived | SD BIOLINE | Serum | 508 | ELISA | 203 | 0 | 0 | 305 | 100 | 100 | 100 | 100 | 100 | 39.96 |
| Loukou [31]2014 | Ivory Coast | Archived | Precise HIV | Serum | 508 | ELISA | 202 | 8 | 1 | 297 | 98.23 | 99.5 | 97.4 | 99.7 | 96.2 | 59.06 |
| Loukou [31]2014 | Ivory Coast | Archived | BCP-2 | Serum | 508 | ELISA | 203 | 1 | 0 | 304 | 99.8 | 100 | 99.7 | 100 | 99.5 | 40.16 |
| Loukou [31]2014 | Ivory Coast | Archived | OraQuick | Serum | 508 | ELISA | 203 | 0 | 0 | 305 | 100 | 100 | 100 | 100 | 100 | 39.96 |
| Loukou [31]2014 | Ivory Coast | Archived | Visiteck | Serum | 508 | ELISA | 202 | 2 | 1 | 303 | 99.41 | 99.5 | 99.3 | 99.7 | 99 | 40.16 |
| Loukou [31]2014 | Ivory Coast | Archived | HIV-Stat | Serum | 508 | ELISA | 203 | 1 | 0 | 304 | 99.8 | 100 | 99.7 | 100 | 99.5 | 40.16 |
| Ondondo [32]2013 | Kenya | HIV serodiscordant couples | SD Bioline/Syphilis | Serum | 680 | ELISA | 345 | 1 | 0 | 352 | 346 | 100 | 100 | 697 | n/a | n/a |
| Bristow[33] 2014 | Kenya | Archived | SD Bioline/Syphilis | Serum | 698 | Determine or Unigold | 345 | 1 | 0 | 352 | 346 | 100 | 100 | 697 | n/a | n/a |

positives (TP), Total Negatives (TN), False Positives (FP) and False Negatives (FN)

## Diagnostic Accuracy of HIV tests

### 1. Quantitative findings

#### Meta-analysis of diagnostic accuracy

**Table 3** shows a meta-analysis of all studies included in this quantitative analysis. The result indicates that the RDT sensitivities were 99% [95% CI=99%-100%], while specificity was optimal at 100% [95% CI=99%-100%]. The Diagnostic odds ratio (DOR) of 44,612 [95% CI=14323-138954], indicates better RDT test performance. The likelihood ratio of a positive test result (LR+) of 317.61 [95% CI=136.61-738.47] was evident.

**Table 3.** Meta-analysis of diagnostic Accuracy of all test Kits.

|  | Coef. | Std. Err. | z | P>z | [95% Conf. | Interval] |
| --- | --- | --- | --- | --- | --- | --- |
| Bivariate |  |  |  |  |  |  |
| E(logitSe) | 4.94 | 0.31 |  |  | 4.33 | 5.55 |
| E(logitSp) | 5.76 | 0.43 |  |  | 4.92 | 6.61 |
| Var(logitSe) | 2.33 | 0.78 |  |  | 1.20 | 4.49 |
| Var(logitSp) | 5.11 | 1.71 |  |  | 2.66 | 9.83 |
| Corr(logits) | 0.32 | 0.20 |  |  | -0.10 | 0.65 |
| HSROC |  |  |  |  |  |  |
| Lambda | 10.75 | 0.58 |  |  | 9.61 | 11.89 |
| Theta | 0.64 | 0.54 |  |  | -0.43 | 1.71 |
| Beta | 0.39 | 0.23 | 1.72 | 0.09 | -0.05 | 0.84 |
| s2alpha | 9.13 | 2.61 |  |  | 5.22 | 16.00 |
| s2theta | 1.16 | 0.45 |  |  | 0.55 | 2.48 |
| Summary Estimates |  |  |  |  |  |  |
| **Sensitivity** | **0.99** | **0.00** |  |  | **0.99** | **1.00** |
| **Specificity** | **1.00** | **0.00** |  |  | **0.99** | **1.00** |
| **DOR** | **44612.33** | **25860.52** |  |  | **14323.11** | **138954.50** |
| **LR+** | **317.61** | **136.73** |  |  | **136.61** | **738.47** |
| LR- | 0.01 | 0.00 |  |  | 0.00 | 0.01 |
| **1/LR-** | **140.46** | **43.64** |  |  | **76.40** | **258.24** |
| **DOR=Diagnostic Odds ratio, LR =Likelihood Ratio (positive or negative)** |  |  |  |  |  |  |

**Table 4** shows a comparison of performance between single tests and dual tests. A single test’s diagnostic odds ratio estimates performed better than a dual test: dual test DOR=44612.33 and single test DOR=14323.11. However, the confidence intervals overlapped, suggesting overall similarity.

**Table 4.**
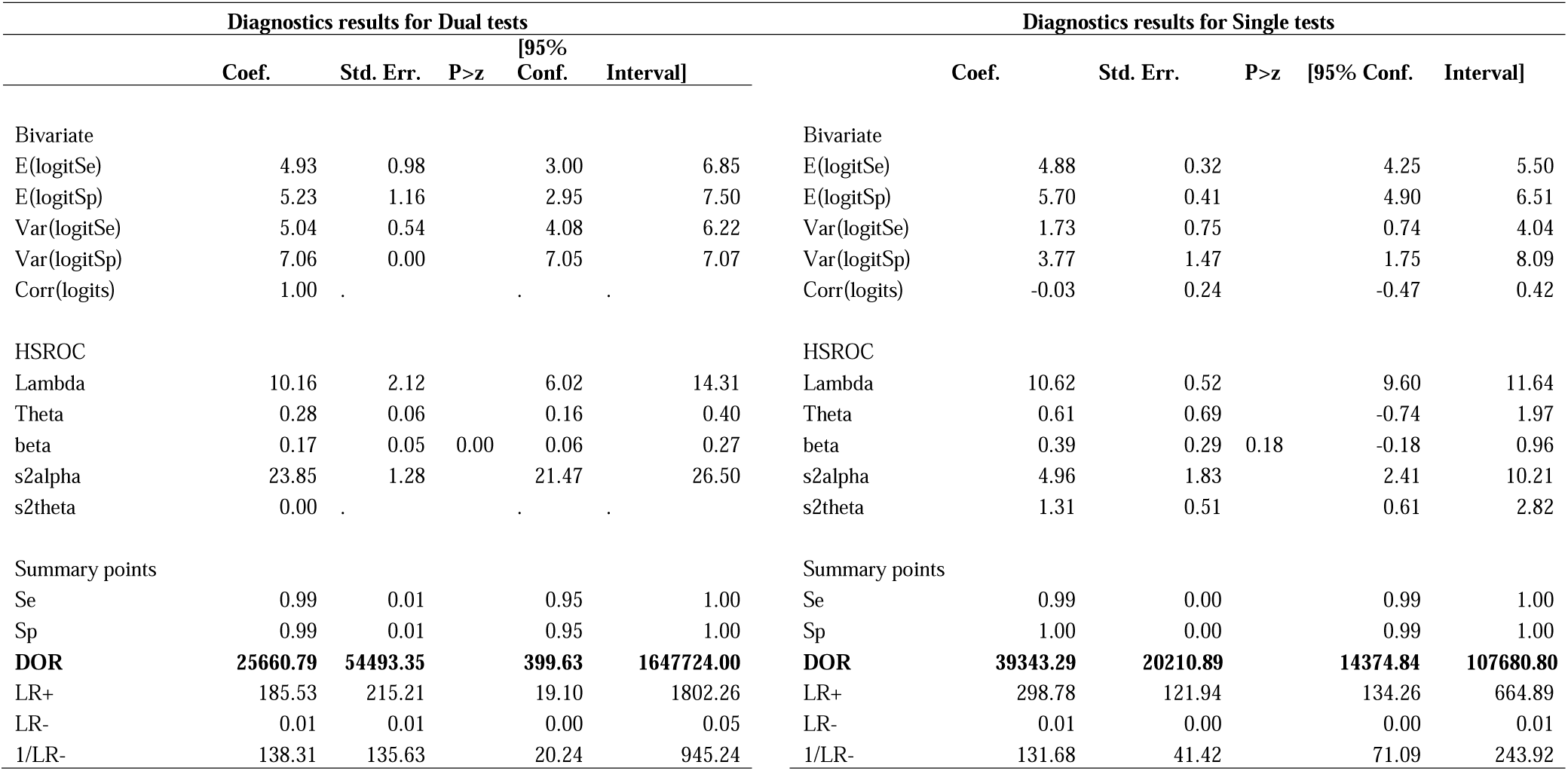
Comparison of diagnostic performance between single and dual tests.

**Table 5a.** Comparison of the performance of RDT samples.

| **RDT sample type** | **variable** | **mean** | **SEM** | **SD** | **min** | **max** |
| --- | --- | --- | --- | --- | --- | --- |
| Blood | DOR | 12359.79 | 7784.036 | 23352.11 | 1.89675 | 72213.9 |
|  | Sensitivity | 85.15887 | 6.670319 | 32.67776 | 0.638 | 100 |
|  | Specificity | 86.51629 | 6.760616 | 33.12012 | 0.518 | 100 |
|  | NPV | 86.675 | 8.374253 | 33.49701 | 0.915 | 100 |
|  | PPV | 84.12 | 8.199338 | 32.79735 | 0.15 | 100 |
| Saliva | DOR | . | . | . | . | . |
|  | Sensitivity | 70.937 | 18.06734 | 47.8017 | 0.972 | 100 |
|  | Specificity | 71.57143 | 18.22166 | 48.20998 | 1 | 100 |
|  | NPV | 79.5794 | 19.65146 | 43.94201 | 0.997 | 100 |
|  | PPV | 80.2 | 19.8 | 44.27415 | 1 | 100 |
| Saliva/Whole blood | DOR | 4424.537 | . | . | 4424.537 | 4424.537 |
|  | Sensitivity | 19.69 | 18.75252 | 41.93191 | 0.872 | 94.7 |
|  | Specificity | 20.716 | 19.721 | 44.0975 | 0.983 | 99.6 |
|  | NPV | 49.9495 | 48.9505 | 69.22646 | 0.999 | 98.9 |
|  | PPV | 49.489 | 48.511 | 68.60491 | 0.978 | 98 |
| Serum | DOR | 19051.13 | 11551.87 | 16336.82 | 7499.25 | 30603 |
|  | Sensitivity | 82.22917 | 11.01803 | 38.16759 | 0.95 | 100 |
|  | Specificity | 91.28333 | 8.210146 | 28.44078 | 1 | 100 |
|  | NPV | 199.4903 | 74.69569 | 247.7376 | 0.993 | 697 |
|  | PPV | 88.18889 | 10.90532 | 32.71595 | 1 | 100 |
| Serum/plasma | DOR | . | . | . | . | . |
|  | Sensitivity | 100 | . | . | 100 | 100 |
|  | Specificity | 98 | . | . | 98 | 98 |
|  | NPV | 100 | . | . | 100 | 100 |
|  | PPV | 96.9 | . | . | 96.9 | 96.9 |
| Serum/plasma/whole blood | DOR | 5151 | . | . | 5151 | 5151 |
|  | Sensitivity | 99.475 | 0.287694 | 0.813722 | 97.8 | 100 |
|  | Specificity | 98.6125 | 0.644326 | 1.82243 | 94.8 | 100 |
|  | NPV | 99.5 | 0.267261 | 0.755929 | 98 | 100 |
|  | PPV | 98.1625 | 1.052537 | 2.977025 | 91.2 | 100 |
| Serum/whole blood | DOR | . | . | . | . | . |
|  | Sensitivity | 99.75 | 0.144338 | 0.288675 | 99.5 | 100 |
|  | Specificity | 97.5 | 1.414214 | 2.828427 | 93.5 | 99.5 |
|  | NPV | . | . | . | . | . |
|  | PPV | . | . | . | . | . |
| Serum | DOR | . | . | . | . | . |
|  | Sensitivity | 98.2 | 1.8 | 3.117691 | 94.6 | 100 |
|  | Specificity | 99.46667 | 0.533333 | 0.92376 | 98.4 | 100 |
|  | NPV | 97.56667 | 2.433333 | 4.214657 | 92.7 | 100 |
|  | PPV | 99.63333 | 0.366667 | 0.635085 | 98.9 | 100 |

**Table 5b.**
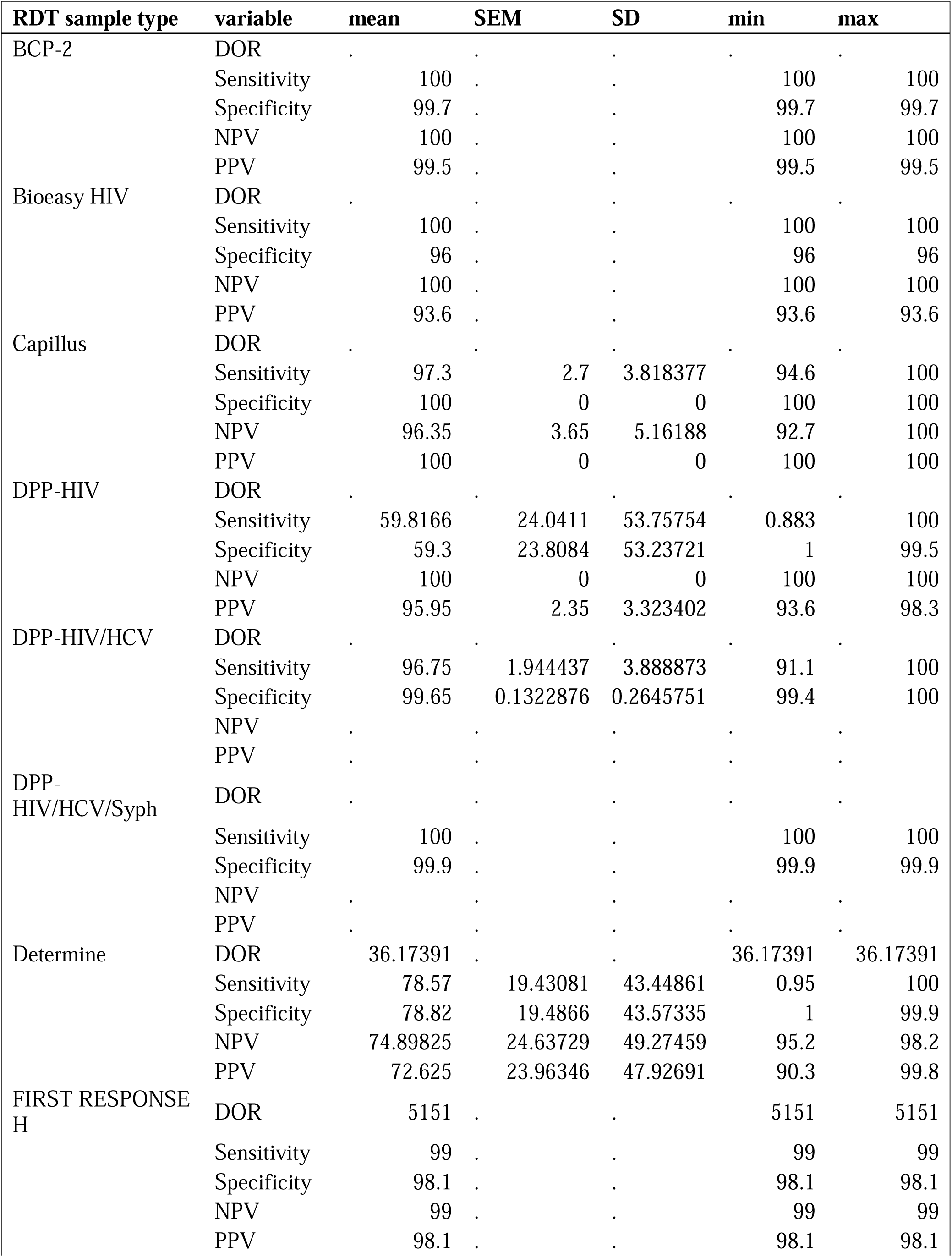
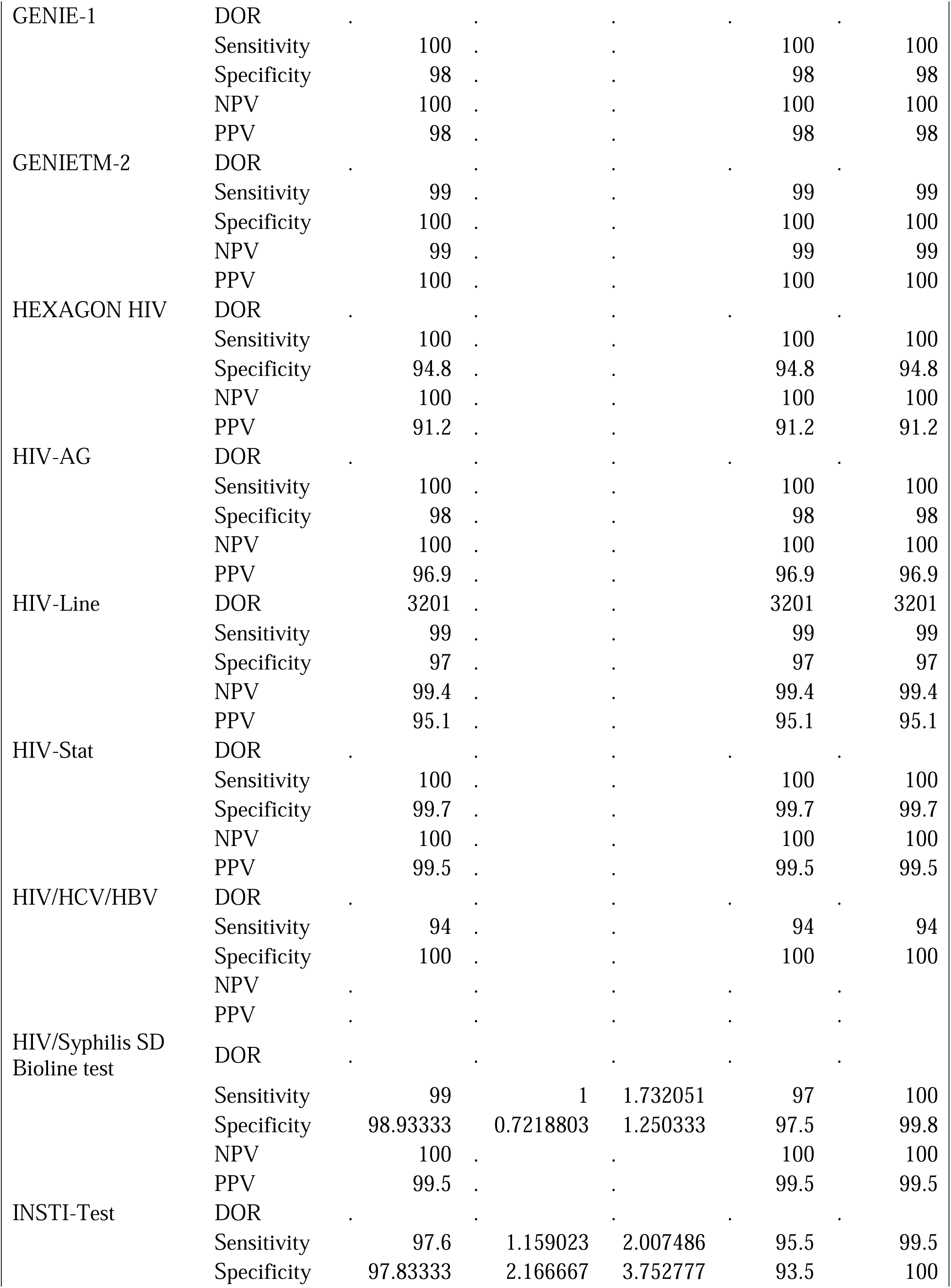
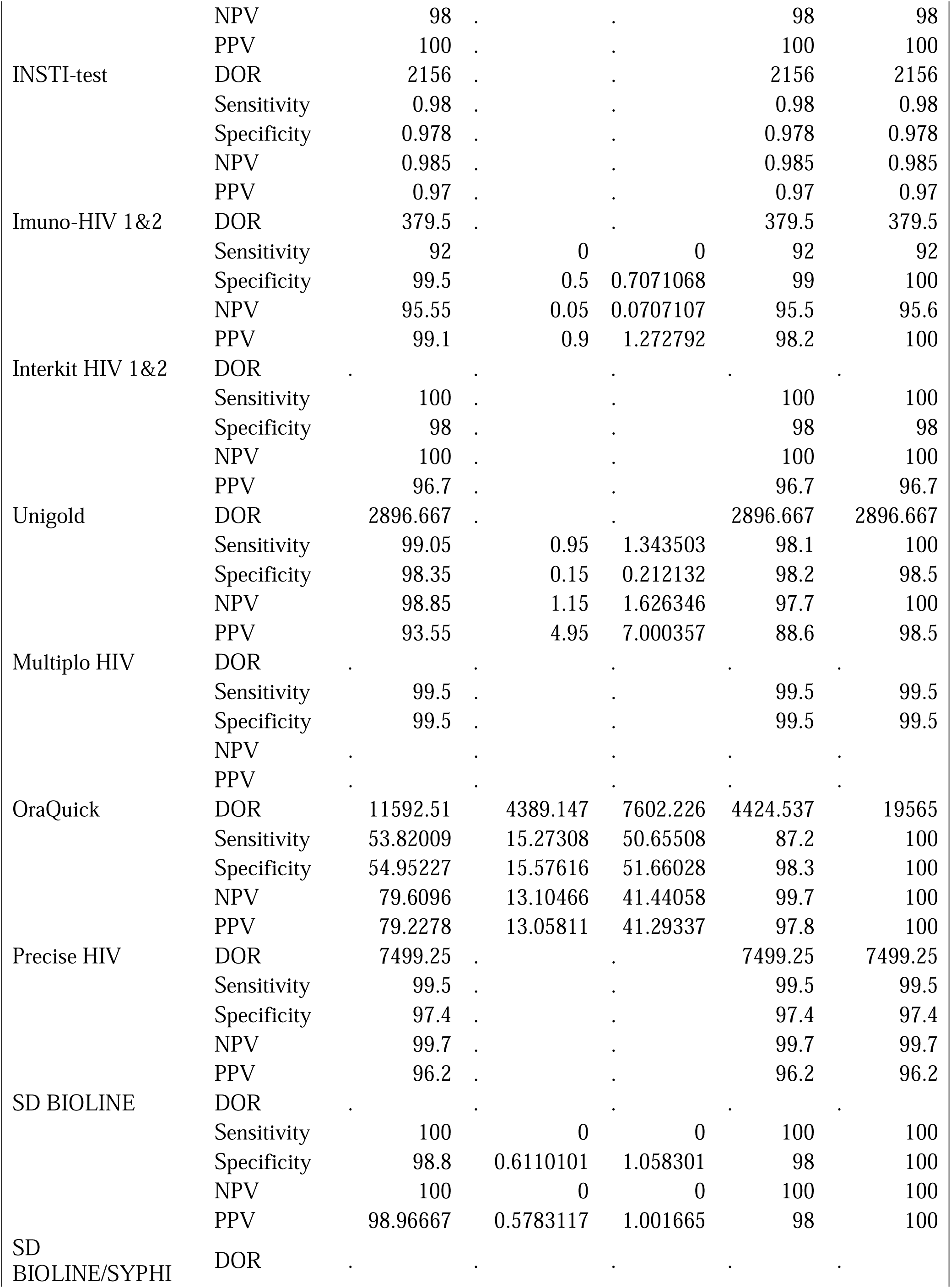
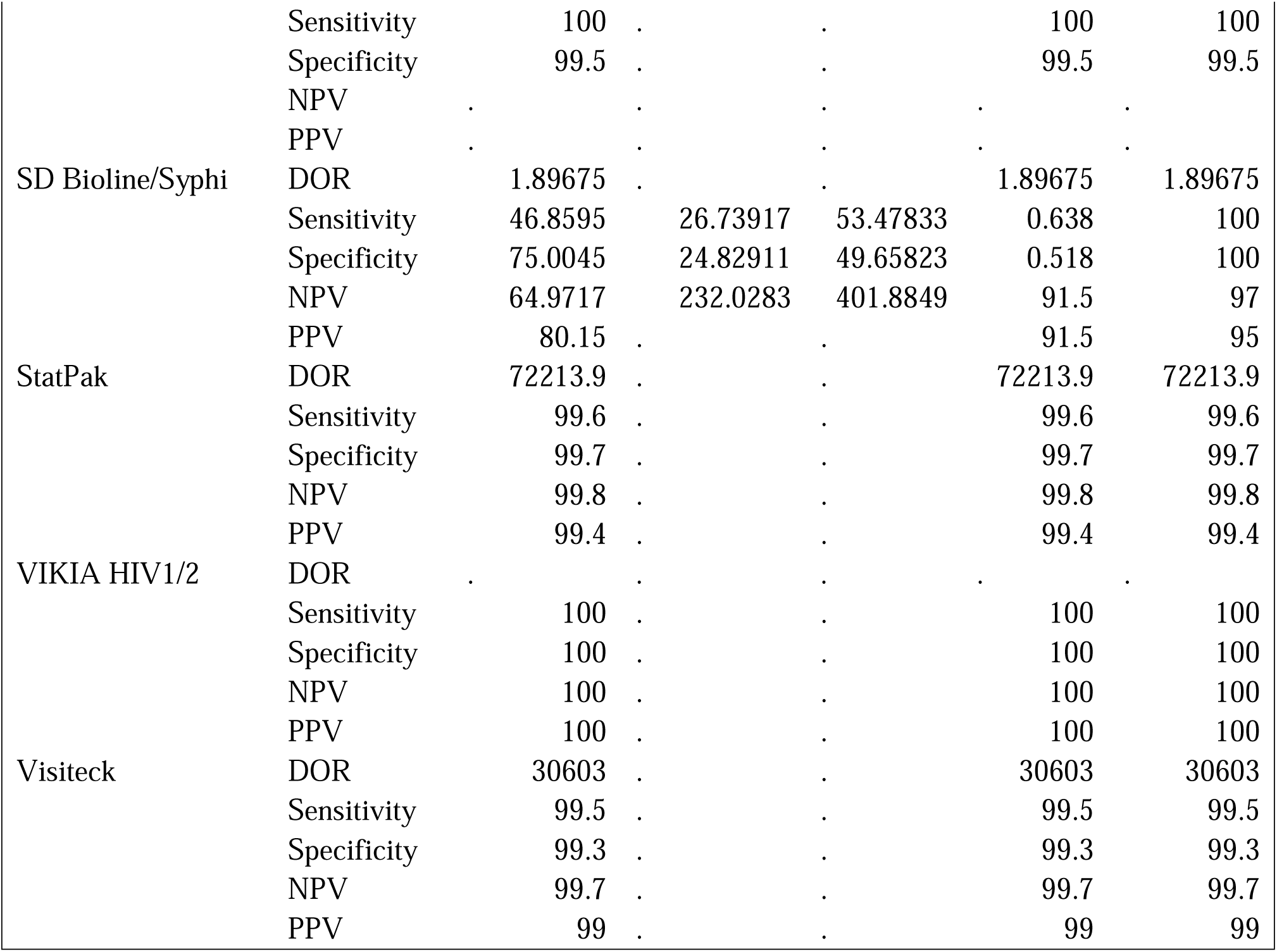
Comparison of individual RDT type.

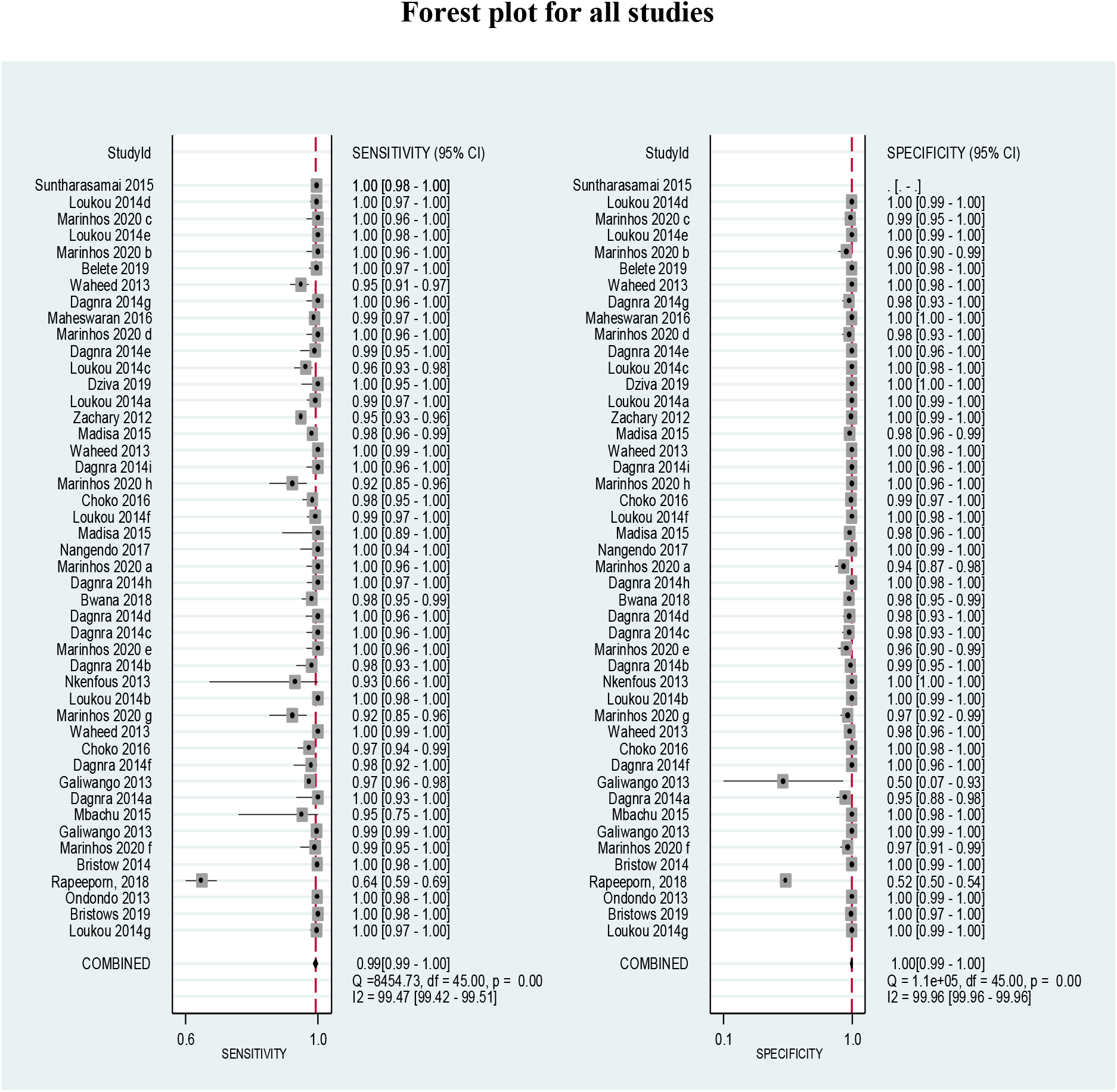

A value of 0% indicates no observed heterogeneity, and values greater than 50% may be considered substantial heterogeneity.

I^2^ for sensitivity was 99.47%, while that for specificity was 99.96, indicating significant heterogeneity justifying stratified analysis.

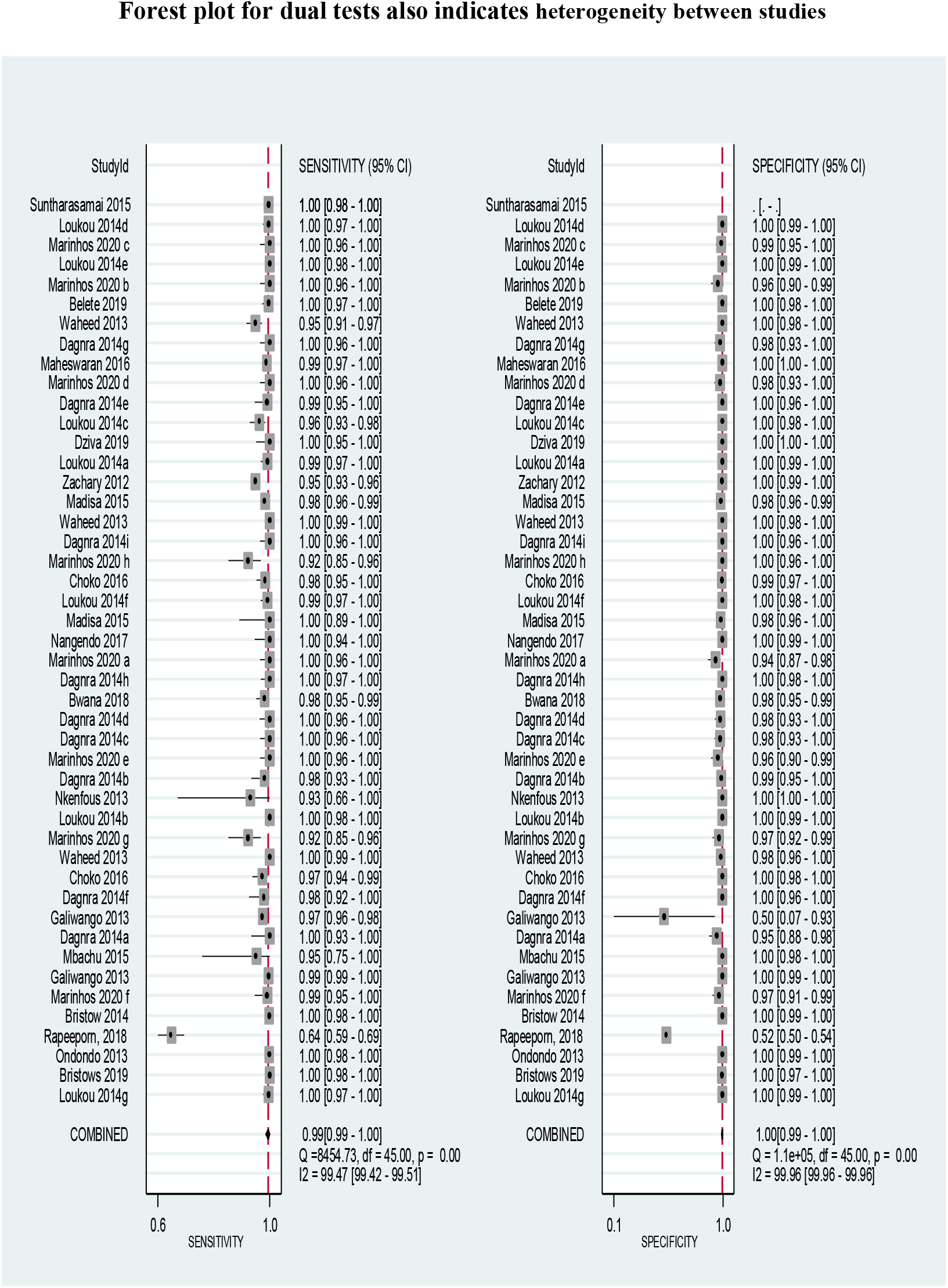

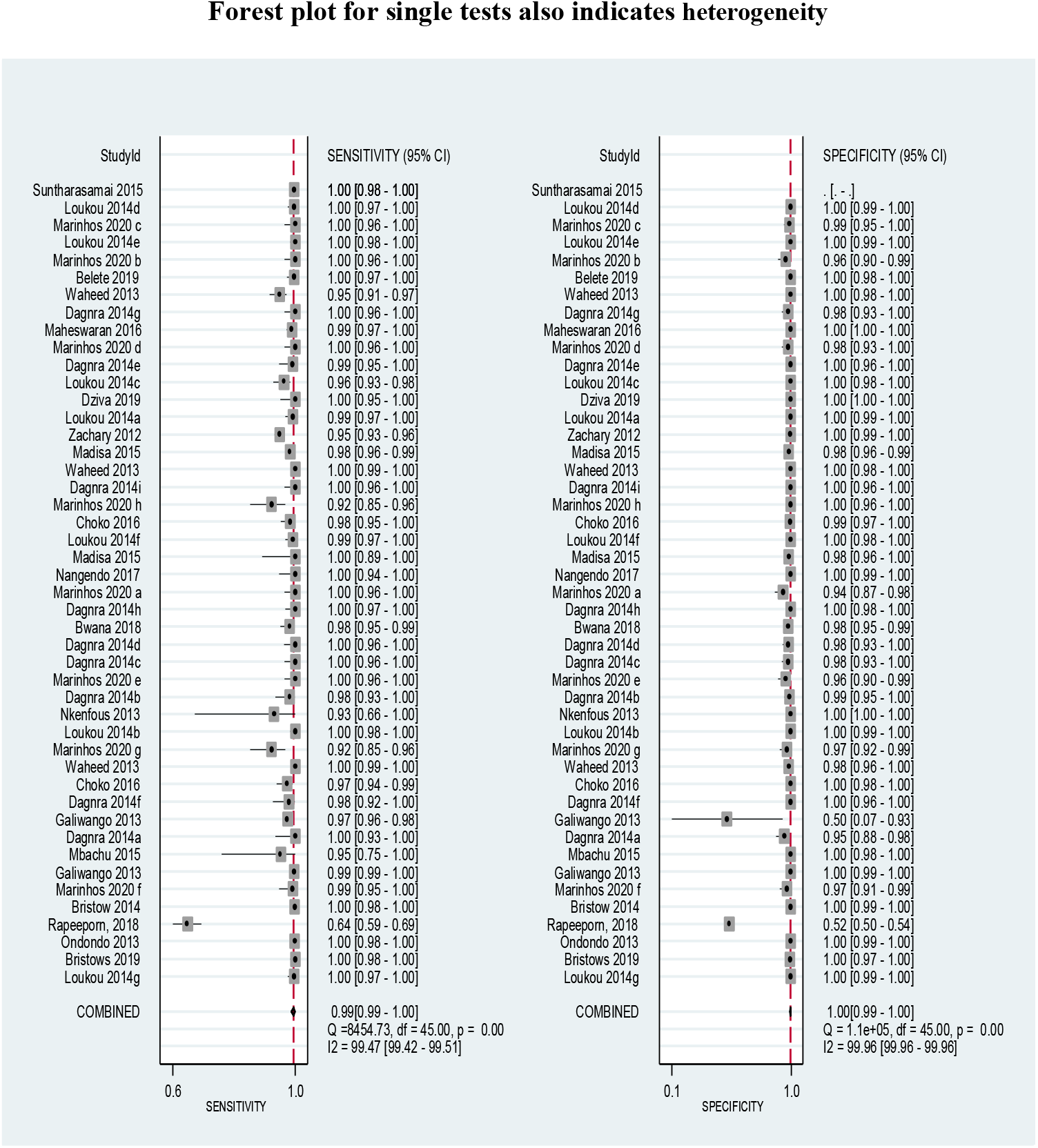

### 2. Qualitative findings

#### a) Acceptability, ease of use for rapid tests for HIV

Overall, most studies showed the acceptability and ease of using diagnostic rapid HIV test kits [34]–[40]. Acceptability analysis papers were all on self-test as they were the ones available. For the HIV self-test Kit, Mugo *et al*. [37] and Kurth *et al*. [39] looked at the uptake, acceptability, and accuracy of oral fluid HIV in a general adult population and found a strong agreement in acceptability and accuracy (>90% accuracy). Frequent testing among index participants (IPs) and their partners – ANC: 51% (27/53), PPC: 68% (62/91), and FSW: 83% (53/64), [35]and 11/18 FSW[38] was documented in three Kenyan studies evaluating the acceptability of self-tests using OraQuick Rapid HIV-1/2 Antibody Test (OraSure Technologies) among FSWs and antenatal and post-partum women in antenatal clinics. In a randomized control trial (RCT) by Masters, Agot [35], participants were more likely to self-test (90.8%, 258/284) in the HIV self-test group than in the comparison group (52%, 148/286). Couples were more likely to test together in the HIV self-test (ST) group than in the comparison group (75.4% versus 33.2%). The majority of the FSWs (89%, 16/18) distributed the self-testing kits to their clients, with each woman distributing them to two or more clients. Most (88%, 14/16) of the clients had positive or neutral reactions during and after using the self-tests [38]. Rumurthy et al. [35] found that most IPs distributed the test kits to their primary partners (ANC: 91% (53/58), PPC: 86% (91/106), and FSW: 75% (64/85)) and other individuals with the 256 IPs with follow-up data distributing 709 self-tests. Virtually all (99%, 442/445) of the self-tests distributed to the implementing partner (IP’s) networks were used by the contacts in the presence of the IP [35].

While exploring the feasibility and acceptability of HIV-testing among PrEP users in Kenya, Ngure *et al*. [41] depicted that 98% of the HIV-uninfected participants who visited the clinics for PrEP refill accepted self-testing using OraQuick, with 93.2% agreeing to have conducted the test at least once. Further, 98% of the participants argued that the OraQuick HIV self-testing kit was easy to use, with 90.8% confirming that they did not require any help to perform the test.

Despite the overall positive feedback regarding ease of use and acceptability, a few studies reported negative experiences among women who tried to introduce the HIVST to their sex partners, thus underscoring the importance of the usability of rapid test kits in different settings [38]. Two women experienced abuse following the self-test distribution; one experienced sexual and verbal abuse (sexual abuse from one client and two clients), and the other verbal abuse from one client in the study.

Strauss *et al*. [41] explored the potential difference in the stated and revealed preference regarding oral-based HIV testing. The study found that the oral-based test for HIV (HST) was well aligned with the preferences of some truck drivers, especially those whose revealed preference was for HST. This evidence shows a strong correlation between stated preference and revealed preference, elucidating acceptability regarding the use of HIVST uptake in different populations [34]. However, the majority preferred to have these tests done in the clinic setting, where they can receive support from a healthcare worker. These findings concur with those from a randomized control trial among truck drivers in Kenya conducted by Ngure *et al*. that partly aimed at determining the HIV testing location preference [41]. The results signify that most participants (76.4%) preferred testing at the clinic, with the preference being 1.5 times higher among those using Oraquick in the choice arm than those using Colloidal Gold in the Standard of Care (SOC).

#### b) Cost-effectiveness for rapid tests for HIV

Costs may be a potential barrier to accessing HIV self-test kits compared to free HTC that the government provides at health facilities. OraQuick, a rapid HIV 1/2 diagnostic HIV test kit that uses saliva to detect HIV, is among the most expensive leading rapid tests, costing around USD 4 in low-resource-limited countries [42]. A few review studies documented the probable costs of rapid HIV tests in different populations and settings. An RCT in a resource-limited country conducted by Eggman *et al*. [43] among adolescents attending the HIV clinic found that the criteria for offering rapid HIV testing and delivering non-rapid test results varied among seven clinics. This study also determined the costs of using a rapid HIV test and observed that using a rapid HIV test costs USD 22 per patient without brief risk-reduction counseling and USD 46 per patient with counseling included. Estimated incremental annual costs per clinic of implementing universal rapid HIV testing varied by whether brief counseling is conducted and by current clinic testing practices, ranging from a savings of USD 19,500 to a cost of USD 40,700 without counseling and a cost of USD 98,000 to USD 153,900 with counseling. A study by Maheswaran *et al*. [44] established that rapid HIV testing at the healthcare facility may be cost-saving but least effective from the health provider perspective using 2010 WHO ART guidelines compared to a combination of facility HTC and HIVST (USD 294.71 per person; 95% credible interval [CrI], 270.79-318.45) and least effective (11.64 quality-adjusted life-years [QALYs] per person; 95% CrI, 11.43-11.86). The cost-effective incremental ratio (ICER) for facility HTC plus HIVST, using 2015 WHO ART guidelines, was USD 253.90 (95% CrI, 201.71-342.02) per QALY gained compared with facility HTC and using 2015 WHO ART guidelines [44]. Other studies in this survey also established lower costs associated with facility HIV rapid tests, including those done by self-testing[13]. However, a study done in Malawi by Maheswaran *et al*. [44], assessing the economic costs and quality of life outcomes of HIV treatment, found lower costs associated with HIVST for ART initiation facility HTC (22.79 versus USD 19.92).

A cluster-randomized trial by Obure *et al*. [45] among pregnant women established costs associated with dual testing of pregnant women for HIV and syphilis in Colombia. They established that the cost per pregnant woman tested and the cost per woman treated for syphilis was USD 10.26 and USD 607.99, respectively, in the single rapid diagnostic test. For dual tests, the cost per pregnant woman tested for HIV and syphilis was USD 15.89, while the cost per woman treated for syphilis was USD 1859.26. The costs per woman tested for HIV and syphilis and those treated for syphilis were lower in Cali than in Bogota across both intervention arms. Health worker costs accounted for the highest costs, while treatment costs comprised <1% of the preventive program [45].

## Discussions

Timely detection and treatment are the key pillars supporting the prevention and control of HIV/AIDS. It is thus important that countries and, more so, resource-limited settings select HIV testing strategies and test kits that enable them to achieve the first UNAIDS 95-95-95 target [45], [46]. RDTs have been used optimally in many settings to achieve widespread testing coverage. This has led to the development of some RDTs with varied sensitivity and specificity results[47], [48]. The selection of RDTs continues to suffer from limited research that evaluates the effectiveness of RDTs in terms of their specificity, sensitivity, and sample types [49]. While striving to address these challenges, we conducted a meta-analysis to evaluate the specificity and sensitivity of blood-based RDTs compared with the WB or ELISA-based assay in terms of pooled sensitivity and specificity by meta-analysis. All the studies reported excellent sensitivity and specificity, with an average RDT sensitivity of 99% [95% CI=99%-100%], while specificity was optimal at 100% [95% CI=99%-100%]. The diagnostic odds ratio was DOR=44612 [95% CI=14323-138954]. Our meta-analysis evaluated the differential performance of single as opposed to dual RDT in terms of the diagnostic odds ratio. The dual test DOR of 44612.33 and single test DOR of 14323.11 evidence the superior diagnostic aspects of a single RDT, although there was an overlap in the confidence interval. This is consistent with Xiaojie et al.’s metanalysis results[49]. These results emphasize using the dual test kit as an alternative backup to the first test kit.

Areas under the SROC curves for the most popular RDT (Capillus HIV-1/HIV-2, Unigold, and Determine HIV-1/2) were above 0.99, with sensitivities above 99.9%. In addition to the most popular RTKs, 15 other kits have been evaluated in 15 studies (see mixed assays in Table 4). The pooled sensitivity, specificity, and area under the SROC curve of these assays were 0.998, 0.991, and 1.00, respectively, which demonstrates that blood-based rapid HIV test has comparable accuracy to WB or ELISA.

Our review suggests that blood-based RDTs have high diagnostic accuracy, with comparable estimates across most test kits as the first test in resource-limited settings. Although the sensitivity and specificity of RDTs reagents both exceed 99.5%, they could be compromised due to unstandardized operations in non-laboratory settings [50], [51]. The sensitivity of RT can be reduced in the absence of a quality assurance and evaluation system [52]. Unstandardized operations may lead to an RT false-negative rate of up to 5.4% [53]. RT test inevitably faces other challenges, such as the inability to recheck the same sample, and relatively low sensitivity for early HIV infection [50]

The main limitation is that statistical comparison between subgroups (i.e., different populations) was not possible due to a lack of data. Additionally, only English language studies were included in this meta-analysis. These could have resulted in a potential reporting bias. Different areas can determine the combinations based on the performance of reagents, costs, HIV prevalence, and risk behaviors of populations. These results depict the need for counselors and clients to understand the limitations of RDTs positive results and the necessity to receive confirmatory tests, especially in low HIV epidemic settings.

## Conclusions

The average of the RDT sensitivities were 99% [95% CI=99%-100%], while specificity was optimal at 100% [95% CI=99%-100%]. The diagnostic odds ratio was DOR=44612 [95% CI=14323-138954], thus indicating better RDT test performance. The performance of single test kits in HIV diagnosis was better than those for dual tests.

Overall, our study indicated that RDTs would function, and the ELISA or WB and RDTs should be accessible and extensively used for HIV diagnosis in line with the WHO recommended test and treatment strategy. Additionally, there are comparable results between the dual and the single test kits.

## Key findings

1. A total of 2,120 studies were identified after rigorous screening. Eventually, 26 studies were included in the RDT performance, and 15 studies were included in the analysis for cost-effectiveness and ease of use.
2. In the meta-analysis of 26 studies, the average RDT sensitivities were 99% [95% CI=99%-100%], while specificity was optimal at 100%; 95% CI=99%-100%. The diagnostic odds ratio was DOR=44612 [95% CI=14323-138954], thus indicating better RDT test performance. The performance of single test kits was better than that for dual tests.
3. While there is significant heterogeneity between studies included in the meta-analysis, there is a need for further assessment of the inclusion criteria among the validation studies.
4. Most of the studies demonstrated a significant high level of acceptability of self-test or initiated tests.
5. Several studies have established that cost remains a barrier to accessing RDT testing for HIV, and time to result is a major factor influencing the test preference.
6. Estimated incremental annual costs per clinic of implementing universal rapid HIV testing varied by whether or not brief counseling is conducted and by current clinic testing practices, ranging from a savings of USD 19,500 to a cost of USD 40,700 without counseling and a cost of USD 98,000 to USD 153,900 with counseling [46].
7. Rapid HIV testing at the healthcare facility may be cost-saving but least effective from the health provider perspective using the 2010 WHO ART guidelines compared to a combination of facility HTC and HIVST (USD 294.71 per person; 95% credible interval [CrI], 270.79-318.45) and least effective (11.64 quality-adjusted life-years [QALYs] per person; 95% CrI, 11.43-11.86).

## Data Availability

All data contained within the article are publicly available?

https://nhrl.nascop.org/home

## Ethics approval and consent to participate

The study was approved by the Amref Health Africa institutional ethics review committee. This was a metanalysis and a systematic review of published literatures.

## Consent for publication

Not applicable

## Availability of data and material

All data contained within the article are publicly available?

## Authors’ contributions

LK conceived the study, collected data, analyzed the data and drafted the manuscript; NP supervised data collection, contributed to data analysis and assisted in drafting and submission of the manuscript. NB, PL, VM.RSM, EN, CA, JO and SM participated in data collection and review of the manuscript. RW and JNK are the overall supervisors and contributed to data analysis, drafting and critical revision of the manuscript. All authors approved the final version of the manuscript.

## Acknowledgments

We acknowledge the Ministry of Health Kenya through the National Public Health Laboratory (NPHL) and the National AIDS and STI Control Program (NASCOP) for facilitating sample collection and allocating staff time to write this manuscript. We thank Dr Bhavna Chohan, Washington State University, for reviewing a draft of this manuscript.

## References

[1] J.R. Paño Pardo, “Rapid Diagnostics and Biomarkers for Antimicrobial Stewardship,” in Antimicrobial Stewardship, 2017. doi: 10.1016/b978-0-12-810477-4.00006-4.

[2] C. At et al., “Accuracy of re-reading HIV rapid tests and the effect of prolonged high temperature,” Topics in Antiviral Medicine, vol. 23. 2015.

[3] I. Olugbenga et al., “Clinic-based evaluation study of the diagnostic accuracy of a dual rapid test for the screening of HIV and syphilis in pregnant women in Nigeria,” PLoS ONE, vol. 13, no. 7, 2018, doi: 10.1371/journal.pone.0198698.

[4] C. S. Kosack et al., “Designing HIV testing algorithms based on 2015 WHO guidelines using data from six sites in sub-Saharan Africa,” Journal of Clinical Microbiology, vol. 55, no. 10, 2017, doi: 10.1128/JCM.00962-17.

[5] A. S. Kravitz Del Solar, B. Parekh, M. O. K. Douglas, D. Edgil, J. Kuritsky, and J. Nkengasong, “A Commitment to HIV Diagnostic Accuracy – a comment on ‘Towards more accurate HIV testing in sub-Saharan Africa: a multi-site evaluation of HIV RDTs and risk factors for false positives’ and ‘HIV misdiagnosis in sub-Saharan Africa: a performance of diagnostic algorithms at six testing sites,’” Journal of the International AIDS Society, vol. 21, no. 8. 2018. doi: 10.1002/jia2.25177.

[6] M. J. Page and D. Moher, “Evaluations of the uptake and impact of the Preferred Reporting Items for Systematic reviews and Meta-Analyses (PRISMA) Statement and extensions: A scoping review,” Systematic Reviews, vol. 6, no. 1, 2017, doi: 10.1186/s13643-017-0663-8.

[7] P. M. Bossuyt et al., “The STARD statement for reporting studies of diagnostic accuracy: Explanation and elaboration,” Clinical Chemistry, vol. 49, no. 1, 2003, doi: 10.1373/49.1.7.

[8] P. F. Whiting et al., “Quadas-2: A revised tool for the quality assessment of diagnostic accuracy studies,” Annals of Internal Medicine, vol. 155, no. 8. 2011. doi: 10.7326/0003-4819-155-8-201110180-00009.

[9] R. M. Harbord and P. Whiting, “Metandi: Meta-analysis of diagnostic accuracy using hierarchical logistic regression,” Stata Journal, vol. 9, no. 2, 2009, doi: 10.1177/1536867x0900900203.

[10] A. S. Glas, J. G. Lijmer, M. H. Prins, G. J. Bonsel, and P. M. M. Bossuyt, “The diagnostic odds ratio: A single indicator of test performance,” Journal of Clinical Epidemiology, vol. 56, no. 11, 2003, doi: 10.1016/S0895-4356(03)00177-X.

[11] P. Bwana, L. Ochieng, and M. Mwau, “Performance and usability evaluation of the INSTI HIV self-test in Kenya for qualitative detection of antibodies to HIV,” PLoS ONE, vol. 13, no. 9, 2018, doi: 10.1371/journal.pone.0202491.

[12] A. T. Choko et al., “Initial accuracy of HIV Rapid test kits stored in suboptimal conditions and validity of delayed reading of oral fluid tests,” PLoS ONE, vol. 11, no. 6, 2016, doi: 10.1371/journal.pone.0158107.

[13] H. Maheswaran et al., “Cost and quality of life analysis of HIV self-testing and facility-based HIV testing and counselling in Blantyre, Malawi,” BMC Medicine, vol. 14, no. 1, 2016, doi: 10.1186/s12916-016-0577-7.

[14] J. Nangendo et al., “Diagnostic accuracy and acceptability of rapid HIV oral testing among adults attending an urban public health facility in Kampala, Uganda,” PLoS ONE, vol. 12, no. 8, 2017, doi: 10.1371/journal.pone.0182050.

[15] C. N. Nkenfou et al., “Evaluation of OraQuick® HIV-1/2 as oral rapid test,” African Journal of Infectious Diseases, vol. 7, no. 2, 2013, doi: 10.4314/ajid.v7i2.2.

[16] M. Jaspard et al., “Finger-stick whole blood HIV-1/-2 home-use tests are more sensitive than oral fluid-based in-home HIV tests,” PLoS ONE, vol. 9, no. 6, 2014, doi: 10.1371/journal.pone.0101148.

[17] I. I. Mbachu et al., “The evaluation of accuracy of serial rapid HIV test algorithm in the diagnosis of HIV antibodies among pregnant women in southeast Nigeria Pregnancy and Childbirth,” BMC Research Notes, vol. 8, no. 1, 2015, doi: 10.1186/s13104-015-1454-8.

[18] P. Suntharasamai et al., “Assessment of oral fluid HIV test performance in an HIV pre-exposure prophylaxis trial in Bangkok, Thailand,” PLoS ONE, vol. 10, no. 12, 2015, doi: 10.1371/journal.pone.0145859.

[19] R. Wongkanya et al., “HIV rapid diagnostic testing by lay providers in a key population-led health service programme in Thailand,” Journal of Virus Eradication, vol. 4, no. 1, 2018, doi: 10.1016/s2055-6640(20)30235-1.

[20] W. Belete et al., “Evaluation of diagnostic performance of noninvasive HIV self-testing kit using oral fluid in Addis Ababa, Ethiopia: A facility-based cross-sectional study,” PLoS ONE, vol. 14, no. 1, 2019, doi: 10.1371/journal.pone.0210866.

[21] C. D. Chikwari et al., “Diagnostic Accuracy of Oral Mucosal Transudate Tests Compared with Blood-Based Rapid Tests for HIV Among Children Aged 18 Months to 18 Years in Kenya and Zimbabwe,” in Journal of Acquired Immune Deficiency Syndromes, 2019, vol. 82, no. 4. doi: 10.1097/QAI.0000000000002146.

[22] D. G. Fisher et al., “Comparisons of New HIV Rapid Test Kit Performance,” AIDS and Behavior, vol. 23, no. 2, 2019, doi: 10.1007/s10461-018-2204-4.

[23] D. Zachary et al., “Field comparison of OraQuick® ADVANCE Rapid HIV-1/2 antibody test and two blood-based rapid HIV antibody tests in Zambia,” BMC Infectious Diseases, vol. 12, 2012, doi: 10.1186/1471-2334-12-183.

[24] R. M. Galiwango et al., “Evaluation of current rapid HIV test algorithms in Rakai, Uganda,” Journal of Virological Methods, vol. 192, no. 1–2, 2013, doi: 10.1016/j.jviromet.2013.04.003.

[25] S. Asiimwe, J. Oloya, X. Song, and C. C. Whalen, “Accuracy of Un-supervised Versus Provider-Supervised Self-administered HIV Testing in Uganda: A Randomized Implementation Trial,” AIDS and Behavior, vol. 18, no. 12, 2014, doi: 10.1007/s10461-014-0765-4.

[26] M. Mine, S. Chishala, K. Makhaola, T. A. Tafuma, J. Bolebantswe, and M. B. Merrigan, “Performance of rapid HIV testing by lay counselors in the field during the behavioral and biological surveillance survey among female sex workers and men who have sex with men in Botswana,” Journal of Acquired Immune Deficiency Syndromes, vol. 68, no. 3, 2015, doi: 10.1097/QAI.0000000000000434.

[27] C. C. Bristow, S. K. V. Rivera, L. B. Ramos Cordova, L. J. Q. Palacios, K. A. Konda, and J. D. Klausner, “Dual rapid test for HIV and syphilis: A laboratory evaluation of the diagnostic accuracy of the Standard Q HIV/Syphilis Combo Test,” Diagnostic Microbiology and Infectious Disease, vol. 94, no. 1, 2019, doi: 10.1016/j.diagmicrobio.2018.11.018.

[28] F. L. de O. Marinho, N. L. de L. Santos, S. P. F. Neves, and L. de S. Vasconcellos, “Performance evaluation of eight rapid tests to detect HIV infection: A comparative study from Brazil,” PLoS One, vol. 15, no. 8, 2020, doi: 10.1371/journal.pone.0237438.

[29] U. Waheed, K. Hayat, B. Ahmad, Y. Waheed, and H. A. Zaheer, “Evaluation of HIV/AIDS diagnostics kits and formulation of a testing strategy for Pakistan,” Journal of Clinical Virology, vol. 56, no. 4, 2013, doi: 10.1016/j.jcv.2012.12.012.

[30] A. Y. Dagnra et al., “Evaluation of 9 rapid diagnostic tests for screening HIV infection, in Lomé, Togo,” Medecine et Maladies Infectieuses, vol. 44, no. 11–12, 2014, doi: 10.1016/j.medmal.2014.10.007.

[31] Y. G. Loukou, M. A. Cabran, Z.N. Yessé, B. M. O. Adouko, S. J. Lathro, and K. B. T. Agbessi-Kouassi, “Performance of rapid tests and algorithms for HIV screening in Abidjan, Ivory Coast,” J Int Assoc Provid AIDS Care, vol. 13, no. 1, 2014, doi: 10.1177/2325957413488168.

[32] R. O. Ondondo, J. B. Odoyo, and E. A. Bukusi, “O15.5 Performance Characteristics of SD Bio Line Rapid HIV-Syphilis Duo Test Kit For Simultaneous Detection of HIV and Syphilis Infections,” Sexually Transmitted Infections, vol. 89, no. Suppl 1, 2013, doi: 10.1136/sextrans-2013-051184.0171.

[33] C. C. Bristow et al., “Multisite laboratory evaluation of a dual human immunodeficiency virus (HIV)/Syphilis point-of-care rapid test for simultaneous detection of HIV and Syphilis infection,” Open Forum Infectious Diseases, vol. 1, no. 1, 2014, doi: 10.1093/ofid/ofu015.

[34] M. Strauss et al., “Stated and revealed preferences for HIV testing: Can oral self-testing help to increase uptake amongst truck drivers in Kenya?,” BMC Public Health, vol. 18, no. 1, 2018, doi: 10.1186/s12889-018-6122-1.

[35] S. H. Masters, K. Agot, B. Obonyo, S. Napierala Mavedzenge, S. Maman, and H. Thirumurthy, “Promoting Partner Testing and Couples Testing through Secondary Distribution of HIV Self-Tests: A Randomized Clinical Trial,” PLoS Medicine, vol. 13, no. 11, 2016, doi: 10.1371/journal.pmed.1002166.

[36] H. Thirumurthy, S. H. Masters, S. N. Mavedzenge, S. Maman, E. Omanga, and K. Agot, “Promoting male partner HIV testing and safer sexual decision making through secondary distribution of self-tests by HIV-negative female sex workers and women receiving antenatal and post-partum care in Kenya: a cohort study,” The Lancet HIV, vol. 3, no. 6, 2016, doi: 10.1016/S2352-3018(16)00041-2.

[37] P. M. Mugo et al., “Uptake and acceptability of oral HIV self-testing among community pharmacy clients in Kenya: A feasibility study,” PLoS ONE, vol. 12, no. 1, 2017, doi: 10.1371/journal.pone.0170868.

[38] S. Maman et al., “A qualitative study of secondary distribution of HIV self-test kits by female sex workers in Kenya,” PLoS ONE, vol. 12, no. 3, 2017, doi: 10.1371/journal.pone.0174629.

[39] A. E. Kurth et al., “Accuracy and Acceptability of Oral Fluid HIV Self-Testing in a General Adult Population in Kenya,” AIDS and Behavior, vol. 20, no. 4, 2016, doi: 10.1007/s10461-015-1213-9.

[40] S. Kalibala, W. Tun, P. Cherutich, A. Nganga, E. Oweya, and P. Oluoch, “Factors associated with acceptability of HIV self-testing among health care workers in Kenya,” AIDS and Behavior, vol. 18, no. SUPPL. 4, 2014, doi: 10.1007/s10461-014-0830-z.

[41] K. Ngure et al., “Feasibility and acceptability of HIV self-testing among pre-exposure prophylaxis users in Kenya,” J Int AIDS Soc, vol. 20, no. 1, 2017, doi: 10.7448/IAS.20.1.21234.

[42] V. Cambiano, S. N. Mavedzenge, and A. Phillips, “Modelling the potential population impact and cost-effectiveness of self-testing for HIV: Evaluation of data requirements,” AIDS and Behavior, vol. 18, no. SUPPL. 4, 2014, doi: 10.1007/s10461-014-0824-x.

[43] A. A. Eggman et al., “The cost of implementing rapid HIV testing in sexually transmitted disease clinics in the United States,” Sexually Transmitted Diseases, vol. 41, no. 9, 2014, doi: 10.1097/OLQ.0000000000000168.

[44] H. Maheswaran et al., “Cost-Effectiveness of Community-based Human Immunodeficiency Virus Self-Testing in Blantyre, Malawi,” Clinical Infectious Diseases, vol. 66, no. 8, 2018, doi: 10.1093/cid/cix983.

[45] C. D. Obure et al., “A comparative analysis of costs of single and dual rapid HIV and syphilis diagnostics: Results from a randomised controlled trial in Colombia,” Sexually Transmitted Infections, vol. 93, no. 7, 2017, doi: 10.1136/sextrans-2016-052961.

[46] Q. Ma et al., “Rapid HIV antibody testing among men who have sex with men who visited a gay bathhouse in Hangzhou, China: A cross-sectional study,” BMJ Open, vol. 5, no. 9, 2015, doi: 10.1136/bmjopen-2015-008661.

[47] W. Urassa, S. Nozohoor, S. Jaffer, K. Karama, F. Mhalu, and G. Biberfeld, “Evaluation of an alternative confirmatory strategy for the diagnosis of HIV infection in Dar Es Salaam, Tanzania, based on simple rapid assays,” Journal of Virological Methods, vol. 100, no. 1–2, 2002, doi: 10.1016/S0166-0934(01)00408-6.

[48] D. Ménard et al., “Evaluation of rapid HIV testing strategies in under equipped laboratories in the Central African Republic,” Journal of Virological Methods, vol. 126, no. 1–2, 2005, doi: 10.1016/j.jviromet.2005.01.023.

[49] J. Pavie et al., “Sensitivity of five rapid HIV tests on oral fluid or Finger-Stick whole blood: A real-time comparison in a healthcare setting,” PLoS ONE, vol. 5, no. 7, 2010, doi: 10.1371/journal.pone.0011581.

[50] R. P. Walensky et al., “Revising expectations from rapid HIV tests in the emergency department,” Annals of Internal Medicine, vol. 149, no. 3, 2008, doi: 10.7326/0003-4819-149-3-200808050-00003.

[51] N. P. Pai et al., “Head-to-head comparison of accuracy of a rapid point-of-care HIV test with oral versus whole-blood specimens: A systematic review and meta-analysis,” The Lancet Infectious Diseases, vol. 12, no. 5, 2012, doi: 10.1016/S1473-3099(11)70368-1.

[52] S. Phillips, T. C. Granade, C. P. Pau, D. Candal, D. J. Hu, and B. S. Parekh, “Diagnosis of human immunodeficiency virus type 1 infection with different subtypes using rapid tests,” Clinical and Diagnostic Laboratory Immunology, vol. 7, no. 4, 2000, doi: 10.1128/CDLI.7.4.698-699.2000.

[53] K. P. Delaney et al., “Evaluation of the performance characteristics of 6 rapid HIV antibody tests,” Clinical Infectious Diseases, vol. 52, no. 2, 2011, doi: 10.1093/cid/ciq068.

